# Chronic Obstructive Pulmonary Disease Does Not Impair Responses to Resistance Training

**DOI:** 10.1101/2021.02.06.21251254

**Authors:** Knut S. Mølmen, Daniel Hammarström, Gunnar S. Falch, Morten Grundtvig, Lise Koll, Marita Hanestadhaugen, Yusuf Khan, Rafi Ahmad, Bente Malerbakken, Tore J. Rødølen, Roger Lien, Bent R. Rønnestad, Truls Raastad, Stian Ellefsen

## Abstract

**Rationale:** Subjects with chronic obstructive pulmonary disease (COPD) are prone to accelerated decay of muscle strength and mass with advancing age. This is mediated by systemic pathophysiologies, which are also believed to impair responses to exercise training, a notion that remains largely unstudied.

**Objectives:** To investigate the presence of impaired training responsiveness in COPD, measured as responses to resistance training compared to healthy participants.

**Methods:** COPD (GOLD grade II-III, n=20, age 69±5) and Healthy (n=58, age 67±4) conducted identical whole-body resistance training interventions, consisting of two weekly, supervised training sessions for 13 weeks. Leg exercises were performed unilaterally, with one leg conducting high-load training (10 repetitions maximum; RM) and the contralateral leg conducting low-load training (30RM).

**Measurements and Main Results:** Measurements included muscle strength (n=7), endurance performance (n=6), muscle mass (n=2), muscle quality, muscle biology (*vastus lateralis*; muscle fiber characteristics, RNA content including transcriptome) and health-related variables (body composition, blood). For core outcome domains, weighted combined factors were calculated from the range of singular assessments.

COPD showed marked improvements in lower-limb muscle strength/mass/quality and lower-limb/whole-body endurance performance, resembling or exceeding those of Healthy, measured as both relative and absolute change terms. This was accompanied by similar changes in muscle biological hallmarks (total RNA/rRNA content↑, muscle fiber cross-sectional area↑, type IIX proportions↓, changes in the mRNA transcriptome). Neither of the core outcome domains were differentially affected by resistance training load.

**Conclusions:** COPD showed marked, unimpaired and hitherto unrecognized responsiveness to resistance training, rejecting the notion of disease-related impairments in training responsiveness.

## Introduction

Chronic obstructive pulmonary disease (COPD) is associated with impaired cardiorespiratory fitness and decreased skeletal muscle mass and strength, leading to reduced levels of daily activity and reduced quality of life.^1,2^ This deterioration is accompanied by systemic co-morbidities such as reduced levels of testosterone,^3^ vitamin D^4,5^ and oxygen saturation levels,^6,7^ and elevated levels of low-grade inflammation,^8^ arguably leaving COPD subjects in a state of anabolic resistance,^9^ resulting in impaired abilities to adapt to exercise training.^10–12^ In particular, these pathophysiologies are believed to impair adaptations to resistance training, which represent the most potent intervention for improving muscle functions, so also for COPD,^13–16^ and for preventing escalation into late-stage morbidities such as pulmonary cachexia.^17^ At present, the presence of anabolic resistance in COPD subjects and its consequences for responses to resistance training remain circumstantial. A mere single study has compared functional and biological adaptations to resistance training between COPD and healthy controls (ISRCTN ID: 22764439).^18–20^

The primary aim of the present study was to compare the effects of 13 weeks of supervised, whole-body resistance training on a wide range of core health and muscle functional and biological characteristics between COPD and healthy control participants (Healthy). The secondary aim was to investigate the effects of high (10 repetitions maximum; RM) and low (30RM) training loads for these adaptations, and to elucidate inherent functional and biological differences between COPD and Healthy.

## Methods

For in-depth description of study protocols and methods, including description of a placebo-controlled vitamin D_3_ supplementation protocol (randomized clinical trial), see Figure 1-2 and clinicaltrial.gov (ClinicalTrials.gov Identifier: NCT02598830). The vitamin D_3_ perspective is covered in detail elsewhere.^21^

**Figure 1.**
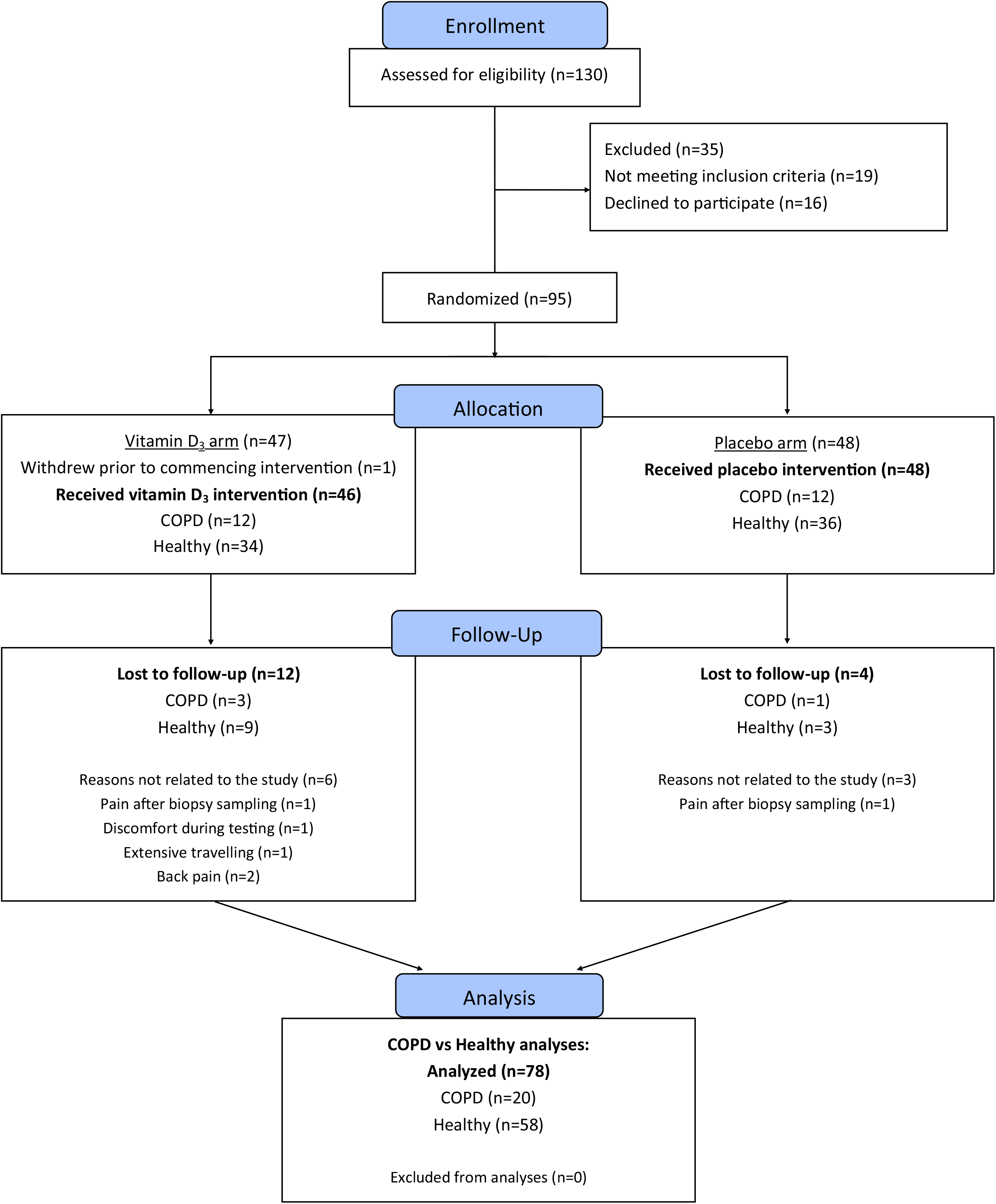
CONSORT flow chart of the study. The study was conducted as a double-blind randomized clinical trial, with the primary aim of investigating the effects of vitamin D_3_ supplementation on resistance training-associated adaptations in a mixed population of older subjects, including both COPD and healthy control subjects (COPD and Healthy, respectively) (ClinicalTrials.gov Identifier: NCT02598830). Vitamin D_3_ supplementation did not affect any primary or secondary outcome, and no conditional effects were observed for COPD vs Healthy in that context.^21^ In the present study, the main purpose was to compare the effects of resistance training between COPD and Healthy participants (COPD, n=20; Healthy, n=58).

### Study ethics and participants

The study was approved by the Regional Committee for Medical and Health Research Ethics (reference no. 2013/1094), preregistered at clinicaltrials.gov (NCT02598830), and conducted according to the Declaration of Helsinki. All participants were informed about the potential risks and discomforts associated with the study and gave their informed consent prior to study enrolment.

Persons with either medical diagnosis of stable COPD (GOLD grade II-III,^22^ predicted FEV_1_ between 80%-30%, FEV_1_/FVC <70% after reversibility testing, n=24, age 70±5) or normal lung function (n=70, age 67±5) were recruited to the study. For study flow chart, see Figure 1. For baseline characteristics, see Table 1.

**Table 1.**
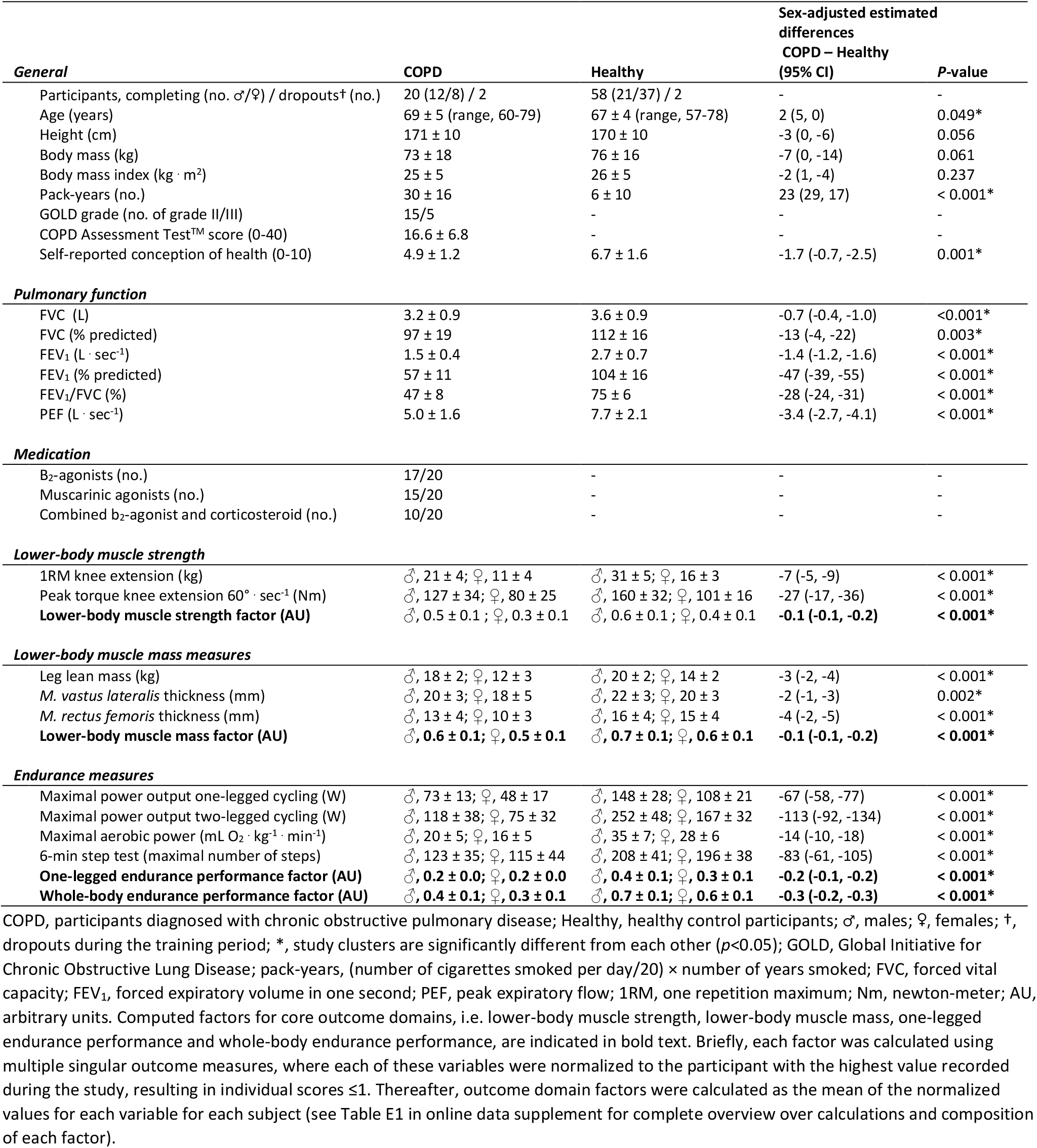
Characteristics of the participants completing the study

### Study conduct

COPD and Healthy conducted identical 13-week resistance training protocols, consisting of two weekly full-body training sessions (Figure 2). Leg exercises were performed unilaterally, with one of the legs of each participant being randomly assigned to perform three sets of 10RM (high-load) and the contralateral leg to perform three sets of 30RM (low-load). All sessions were supervised by qualified personnel. The effectiveness of the training intervention was assessed as a wide range of outcome measures (Figure 2), including multiple assessments of endurance performance, muscle strength and mass, measures of work economy/efficiency, and collection of blood and *vastus lateralis* biopsies (both legs) (Figure 2).

**Figure 2.**
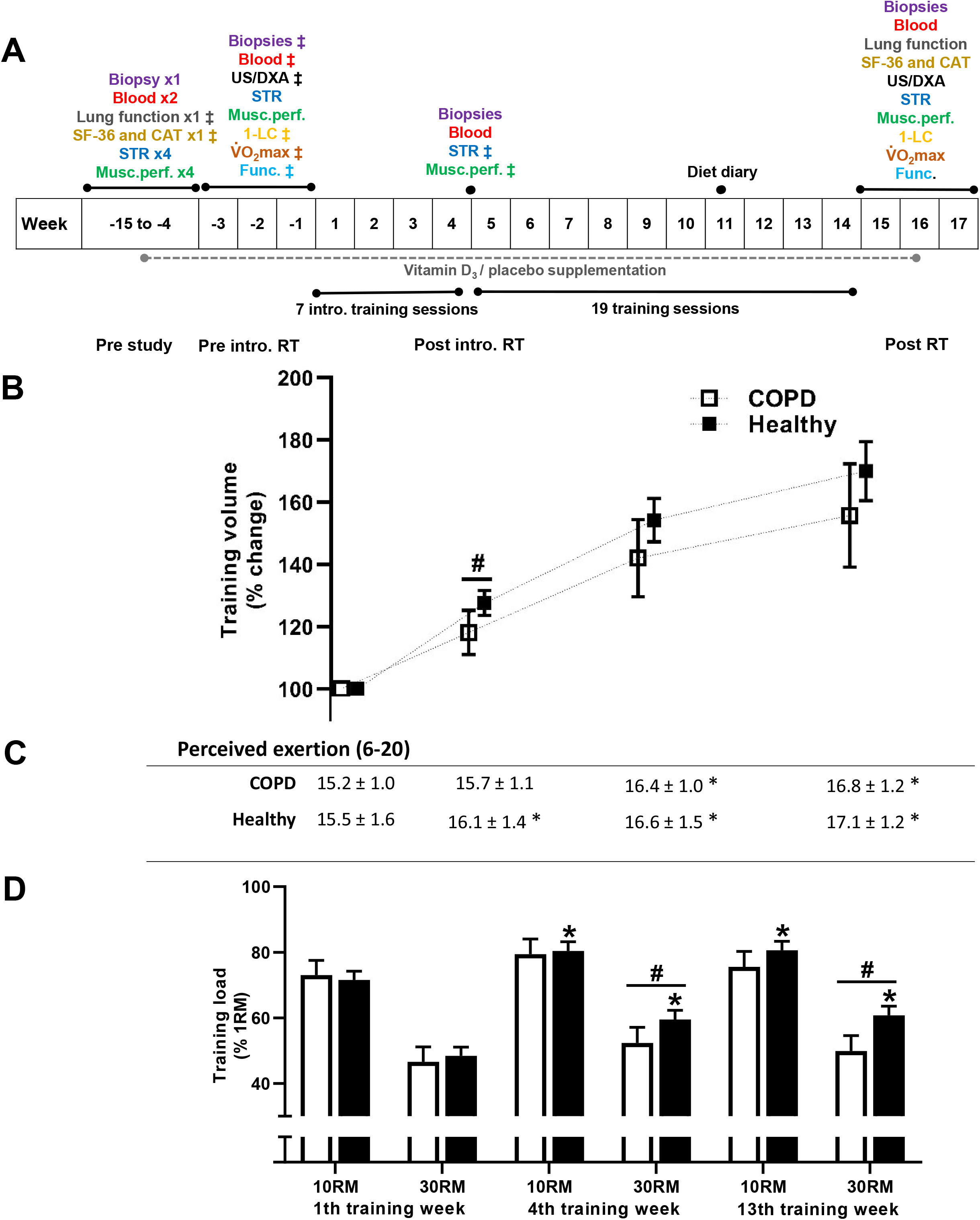
Schematic overview of the study protocol, including its time line (a; ‡ indicates the defined baseline measurement for the specific outcome measure), training volumes during the resistance training (RT) intervention (b), perceived exertion (Borg RPE, 6-20) reported after training sessions (c), and relative training loads (% of 1RM) during the training period (d). Training volume is presented as average increases in per-session for lower-body appendices from the first week of training (kg. repetitions; high-load (10RM) and low-load (30RM) leg press and knee extension combined). COPD, participants diagnosed with chronic obstructive pulmonary disease; Healthy, healthy control participants; *, statistical different from 1th training week; #, statistical difference between COPD and Healthy. Data are presented as means with 95% confidence limits. Methodological notes on retrieval of outcome measures: **i)** Lung function. Spirometry testing was performed following the guidelines from the American Thoracic Society and the European Respiratory Society.^47^ Participants with COPD were tested before and after inhalation of two bronchodilators (salbutamol/ipratropiumbromid). **ii)** Muscle strength and performance (STR and Musc. perf). Muscle strength was assessed as one-repetition maximum (1RM) in unilateral knee extension and leg press, bilateral chest press, and handgrip. Muscle performance was defined as the number of repetitions achieved at 50% of pre-study 1RM and was assessed using unilateral knee extension and bilateral chest press. Isokinetic unilateral knee-extension torque was tested at three angular speeds (60°, 120° and 240°. sec^-1^; Humac Norm, CSMi, Stoughton, MA, USA). **iii)** One-legged cycling and bicycling performance (1-LC and VO_2_max). Participants conducted one-legged cycling tests (Excalibur Sport, Lode BV, Groningen, the Netherlands) to assess O_2_-costs and mechanical efficiency^48^ during submaximal cycling, and maximal one-legged oxygen consumption 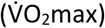 and maximal workload. Maximal two-legged cycling 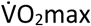 and workload were tested on a separate day. Oxygen consumption was measured using the JAEGER Oxycon Pro^™^ system (Carefusion GmbH, Höchberg, Germany). **iv)** Functional performance (Func.). Functional tests were conducted as the maximal number of sit-to-stands during one minute (seat height: 45 cm) and as the number of steps onto a 20 cm step box during 6 minutes. **v)** Health-related quality of life (SF-36 and CAT). All participants completed the Short Form (36-item) Health Survey (SF-36). COPD participants also completed the COPD Assessment Test (CAT) questionnaire. **vi)** Muscle thickness and body mass composition (US/DXA). Muscle thickness of *m. vastus lateralis* and *m. rectus femoris* were measured using B-mode ultrasonography (SmartUs EXT-1M, Telemed, Vilnius, Lithuania). Body mass composition was measured using dual-energy X-ray absorptiometry (DXA; Lunar Prodigy, GE Healthcare, Madison, WI, USA).

### Blood and muscle measurements

Prior to collection of blood and muscle biopsies, participants were instructed to attend an overnight fast and to avoid heavy physical activity for the last 48 h. Blood samples were analyzed for serum concentrations of hormones, lipids, and markers of iron metabolism and tissue damage, as previously described.^21^ Muscle biopsies were analyzed for muscle fiber type proportions, myonuclei content, muscle fiber cross-sectional area (CSA), and rRNA and mRNA content (total RNA, rRNA subspecies, myosin heavy chain isoforms I, IIA and IIX, and whole-genome transcriptome), as previously described.^21,23,24^ Transcriptome analysis was restricted to a subset of participants (COPD, n=19; Healthy, n=34).

### Data analyses and statistics

For an in-depth description of data analyses and statistics, see the online data supplement. For continuous and non-continuous variables, respectively, linear mixed-effects models and generalized linear mixed-effects models were used to examine differences between COPD and Healthy, both at baseline and as responses to resistance training.

To achieve reliable assessment of core outcome domains, and thus to lower the risk of statistical errors, combined factors were calculated for outcome measures relating to lower-body muscle strength, lower-body muscle mass, one-legged endurance performance and whole-body endurance performance, as previously described.^21^ For details, see online data supplement, Table E1.

For transcriptome analyses, genes were regarded as differentially expressed when the absolute log_2_ fold-change/difference was greater than 0.5 and the adjusted *p*-value (false discovery rate adjusted *per* model coefficient) was below 5%.^23^ Enrichment analyses were performed using two approaches, the non-parametric rank test (Rank) and the directional gene set enrichment analysis (GSEA), where consensus results of those two analyses were interpreted as having larger biological meaning.

Statistical significance was set to *p*<0.05. In both text and figures, data are presented as adjusted marginal means, with or without 95% confidence intervals, unless otherwise stated.

## Results and discussion

### Baseline characteristics: COPD vs Healthy

#### Exercise capacity, body composition and muscle and blood biology

At baseline, COPD displayed impaired exercise capacity compared to Healthy, as expected from previous studies.^2,18,20,25^ This was evident as impaired whole-body performance (range: −41% to −54%, online data supplement, Table E2), and lower-body unilateral muscle strength and endurance performance (−17% to −30%, online data supplement, Table E2), reflecting the cardiorespiratory and muscular limitations inherent to the condition.^26^ In accordance with this, COPD had less lean body mass than Healthy (Δ-13%, online data supplement, Table E2), with 45% of COPD showing signs of sarcopenia, as defined by Baumgartner *et al*.^27^ This difference was unlikely to be caused by the miniscule age difference between COPD and Healthy (Table 1), as this would have implied an annual loss of ∼2.6 kg lean mass *per* year, markedly deviating from the expected loss in this age group (∼0.5 kg *per* year).^28^ The negative effects of COPD for muscle mass was underlined by −9%/-24% smaller *vastus lateralis/rectus femoris* thicknesses (Table 1), corresponding well with difference in leg-specific lean mass (−16%; Table 1), offering potential explanations for the impaired maximal leg muscle strength.

There were also inherent differences between the two study clusters at the muscle biological level, COPD displayed greater proportions of type IIA and IIX muscle fibers in *vastus lateralis* compared to Healthy (32%/23% vs 13%/9%, respectively), with concomitantly lower proportions of type I fibers, corroborating with previous studies.^29,30^ COPD also showed larger CSA (12%, online data supplement, Table E2) and greater myonuclear domain (CSA *per* myonuclei) of type I fibers (Δ20%, online data supplement, Table E2), with no difference being observed for type II fibers. This contrasts previous studies, who have reported smaller or similar CSA in type I fibers in COPD compared to Healthy,^25,31,32^ but may point to a compensatory mechanism for the likely loss of motor units in COPD subjects,^33^ whereby reduced quantities of muscle fibers are compensated for by increased sizes of remaining fibers, as previously reported in rodents.^34^ Furthermore, COPD also affected whole-genome transcriptome profiles and displayed differential expression of 227 genes compared to Healthy (151↑ and 76↓; Figure 3a, online data supplement, Table E3). Hallmark enrichment analysis revealed lower expression of genes involved in *oxidative phosphorylation* (consensus), corroborating with the lower type I proportion, and greater expression of genes involved in regulation of *myogenesis* (Rank) (Figure 3a-b, Table 2; findings confirmed in gene ontology analysis, online data supplement, Table E4), which may be related to the pathophysiological elevation of protein turnover in COPD.^35,36^

**Table 2.** Comparison of Hallmark gene sets identified in whole-genome transcriptome data between COPD and Healthy, assessed at baseline and as resistance training-associated changes.

| Comparison | Gene set | Significance category* | Set size<sup>†</sup> | Rank P-value<sup>‡</sup> | % MSD > 0<sup>§</sup> | GSEA P-value | NES | LE** | Log<sub>2</sub> fold difference in LE (95% CI) |
| --- | --- | --- | --- | --- | --- | --- | --- | --- | --- |
| Baseline: COPD vs. Healthy | Oxidative phosphorylation | Consensus | 190 (200) | 0.007 | 36.8% | <0.001 | -2.10 | 70 (94.3%) | -0.24 (-0.45, -0.13) |
|  | Myogenesis | Rank | 163 (200) | <0.001 | 33.7% | 0.417 | 1.21 | 45 (75.6%) | 0.46 (0.19, 1.5) |
| 3½ weeks training: ΔCOPD vs ΔHealthy | Allograft rejection | GSEA | 115 (200) | 0.956 | 7.8% | 0.014 | 1.71 | 20 (35%) | 0.39 (0.13, 0.76) |
|  | Oxidative phosphorylation | GSEA | 190 (200) | 0.999 | 1.1% | 0.009 | 1.69 | 83 (2.4%) | 0.11 (0.05, 0.39) |
|  | Pancreas beta cells | GSEA | 15 (40) | 0.969 | 6.7% | 0.028 | 1.71 | 3 (33.3%) | 0.35 (0.08, 0.54) |
| Post-RT (13 weeks training): ΔCOPD vs ΔHealthy | Myogenesis | Consensus | 163 (200) | <0.001 | 42.3% | <0.001 | -1.52 | 68 (85.3%) | -0.5 (-1.13, -0.26) |

\* Consensus significance indicates agreement between directional (GSEA) and non-directional (Rank) hypothesis test of overrepresentation (see methods for details). <sup>†</sup> Indicates number of identified genes in the gene set and total number of genes in the gene set in parentheses. <sup>‡</sup> Rank-based enrichment test, based on minimum significant difference (MSD), identifies gene sets that are overrepresented among top-ranked genes without a directional hypothesis. <sup>§</sup> Fraction of genes in gene set with unadjusted 95% CI not spanning zero, i.e. MSD > 0. <sup>||</sup> Gene-set enrichment analysis (GSEA) tests for overrepresentation among top and bottom genes based on Log<sub>2</sub> fold differences or changes $\times -\log_{10}(P\text{-values})$ in comparing differences at baseline or changes from baseline between COPD and Healthy. A positive normalized enrichment score (NES) indicate gene set with higher expression in COPD than Healthy; negative NES indicate gene set with lower expression at respective time-points. \*\* Number of genes in leading edge (LE, genes that contributes to the enrichment score) with the fraction of leading edge genes with unadjusted 95% CI not spanning zero. Δ, change score.

**Figure 3.**
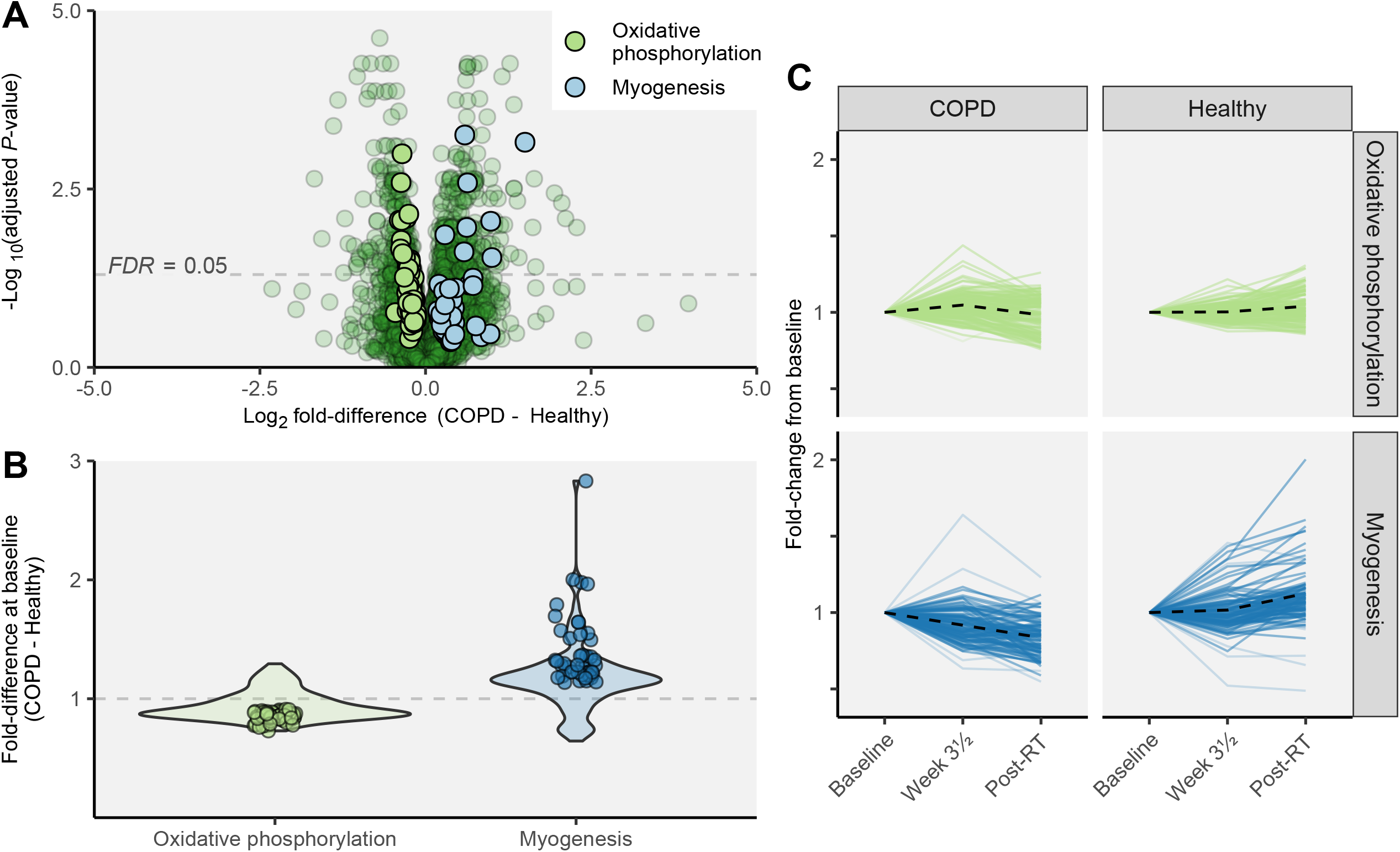
Whole-genome transcriptome analyses of *m. vastus lateralis* in COPD and Healthy. At baseline, numerous genes were differentially expressed between COPD and Healthy. In (a), differences in gene expression between COPD and Healthy are presented with leading edge genes (i.e. genes that contributes to the enrichment score) from two gene sets identified as differentially expressed between COPD and Healthy from gene enrichment analyses (*oxidative phosphorylation* and *myogenesis*; see Table 2). In (b), average fold differences (COPD - Healthy) of genes contributing to baseline differences in *oxidative phosphorylation* and *myogenesis* gene sets are shown as individual data points, and violin plots shows the distribution of all leading edge genes from each gene set. (c) displays the average development of each gene set over time, where the dotted line indicates the mean fold change of all genes contributing to the differential change over time between COPD and Healthy. COPD displayed larger increases in expression of genes relating to *oxidative phosphorylation* after 3½ weeks of training, and more pronounced decreases in genes associated with *myogenesis* to after the training intervention (Post-RT; see Table 2). FDR, false discovery rate-adjusted *p*-value.

For other muscle characteristics, such as the content of total RNA and rRNA *per* amount of muscle tissue, hormonal status in blood (e.g. testosterone) and nutritional status (e.g. protein intake complying with current guidelines^37^), no differences were observed between COPD and Healthy at baseline (online data supplement, Table E2, Table 3). Low-grade inflammation, measured as c-reactive protein levels in blood, was elevated in COPD compared to Healthy at pre-study (5.0 vs 1.6 mg.L^-1^) and tended to differ at baseline (p=0.053; Table 3), as expected from previous studies.^8^

**Table 3.** Effects of the training intervention on body composition and blood variables in COPD and Healthy, assessed as changes from baseline to after completion of the study (per study cluster) and as differential changes between study cluster.

|  | COPD |  |  | Healthy |  |  | Δ COPD vs Δ Healthy |
| --- | --- | --- | --- | --- | --- | --- | --- |
|  | Baseline | Post RT | Time effect (P<0.05) | Baseline | Post RT | Time effect (P<0.05) | (P value) |
| **Dual-energy x-ray absorptiometry** |  |  |  |  |  |  |  |
| Whole-body bone mineral density (g · cm<sup>2</sup>) | 1.13 ± 0.21 | 1.13 ± 0.21 | No | 1.15 ± 0.16 | 1.14 ± 0.15 | No | 0.119 |
| Total lean mass (kg) | 46.7 ± 9.9 | 47.6 ± 10.2 | Yes ↑ | 48.1 ± 10.0 | 48.6 ± 10.0 | Yes ↑ | 0.395 |
| Appendicular lean mass (kg) | 20.3 ± 5.3 | 20.9 ± 5.5 | Yes ↑ | 21.6 ± 5.0 | 21.9 ± 5.0 | Yes ↑ | 0.166 |
| Total fat mass (kg) | 26.4 ± 11.7 | 26.3 ± 11.5 | No | 25.3 ± 9.3 | 24.4 ± 9.2 | Yes ↓ | 0.068 |
| Visceral fat (kg) * | 1.59 ± 1.18 | 1.56 ± 1.21 | No | 1.12 ± 0.98 | 1.01 ± 0.81 | Yes ↓ | 0.138 |
| **Inflammation** |  |  |  |  |  |  |  |
| C-reactive protein (mg · L<sup>-1</sup>) | 3.4 ± 5.0 | 3.6 ± 4.0 | No | 1.7 ± 2.5 | 1.8 ± 3.5 | No | 0.934 |
| **Hormones** |  |  |  |  |  |  |  |
| Cortisol (nmol · L<sup>-1</sup>) * | 307 ± 130 | 310 ± 109 | No | 369 ± 88 | 372 ± 99 | No | 0.861 |
| Growth hormone (μg · L<sup>-1</sup>) | 1.4 ± 2.8 | 1.4 ± 3.1 | No | 1.1 ± 1.7 | 1.3 ± 1.6 | No | 0.837 |
| IGF-1 (nmol · L<sup>-1</sup>) | 15.7 ± 4.2 | 15.0 ± 4.5 | No | 14.4 ± 3.2 | 13.6 ± 3.1 | Yes ↓ | 0.977 |
| Testosterone (nmol · L<sup>-1</sup>)† | 11.2 ± 4.4 | 11.4 ± 4.2 | No | 11.9 ± 3.3 | 12.4 ± 4.2 | No | 0.938 |
| Sex-hormone binding globulin (nmol · L<sup>-1</sup>) | 60 ± 33 | 60 ± 34 | No | 60 ± 22 | 60 ± 21 | No | 0.488 |
| Androstenedione (nmol · L<sup>-1</sup>) | 3.3 ± 2.4 | 3.3 ± 2.4 | No | 3.8 ± 2.7 | 3.8 ± 2.4 | No | 0.984 |
| Parathyroid hormone (pmol · L<sup>-1</sup>) | 5.7 ± 2.6 | 6.0 ± 3.3 | No | 5.0 ± 2.2 | 5.2 ± 1.9 | No | 0.870 |
| **Lipid profile variables** |  |  |  |  |  |  |  |
| Triglycerides (mmol · L<sup>-1</sup>) | 1.2 ± 0.5 | 1.1 ± 0.5 | No | 1.2 ± 0.5 | 1.1 ± 0.6 | Yes ↓ | 0.661 |
| HDL (mmol · L<sup>-1</sup>) | 1.6 ± 0.6 | 1.7 ± 0.5 | No | 1.7 ± 0.5 | 1.7 ± 0.5 | No | 0.523 |
| LDL (mmol · L<sup>-1</sup>) * | 2.8 ± 1.0 | 2.8 ± 1.0 | No | 3.4 ± 1.0 | 3.3 ± 0.8 | No | 0.775 |
| **Iron biology variables** |  |  |  |  |  |  |  |
| Fe<sup>2+</sup> (μmol L<sup>-1</sup>) | 18 ± 7 | 18 ± 6 | No | 18 ± 6 | 18 ± 5 | No | 0.410 |
| Transferrin (g · L<sup>-1</sup>) * | 2.66 ± 0.44 | 2.67 ± 0.45 | No | 2.41 ± 0.27 | 2.38 ± 0.29 | No | 0.563 |
| Ferritin (μg · L<sup>-1</sup>) | 113 ± 92 | 90 ± 81 | Yes ↓ | 139 ± 79 | 133 ± 68 | No | 0.089 |
| **Calcium status** |  |  |  |  |  |  |  |
| Calcium (mmol · L<sup>-1</sup>) | 2.4 ± 0.1 | 2.4 ± 0.1 | No | 2.4 ± 0.1 | 2.4 ± 0.1 | No | 0.865 |
| Albumin-corrected calcium (mmol · L<sup>-1</sup>) | 2.3 ± 0.1 | 2.3 ± 0.1 | No | 2.3 ± 0.1 | 2.3 ± 0.1 | No | 0.802 |
| **Tissue damage variables** |  |  |  |  |  |  |  |
| Aspartate transaminase (units · L<sup>-1</sup>) | 27 ± 9 | 24 ± 6 | No | 26 ± 21 | 26 ± 7 | No | 0.807 |
| Creatine kinase (units · L<sup>-1</sup>) | 112 ± 69 | 123 ± 71 | No | 95 ± 47 | 125 ± 72 | Yes ↑ | 0.523 |

\*, significant difference between COPD and Healthy at baseline; †, only men were included in testosterone analysis; ↓, significant decrease from baseline to post RT (after 13 weeks of resistance training); ↑, significant increase from baseline to post RT. Alpha level at $p < 0.05$ .
Data are presented as means with standard deviations.

### The efficacy of the resistance training intervention: COPD vs Healthy

For both COPD and Healthy, the training intervention was associated with low drop-out rates, high adherence to the protocol, and progressive increases in training volume and muscle strength (for details on these perspectives, see online data supplement). The vitamin D_3_ supplementation RCT of the project did not enhance or affect training-associated changes for any of the primary or secondary outcome measures.^21^

#### Muscle strength, muscle mass, muscle quality and one-legged endurance performance

Overall, COPD showed larger training-associated increases in lower-body muscle strength and mass compared to Healthy (the two legs/training modalities combined), measured as relative changes in combined factors from baseline (Figure 4a), with no difference being observed for absolute changes (Figure 4a). COPD and Healthy showed similarly scaled improvements in muscle quality and one-legged endurance performance (Figure 4a). Notably, neither of these four core outcome domains were differentially affected by resistance training load (neither in COPD nor in Healthy), suggesting that 30RM training is an effective alternative to 10RM training in older individuals (Figure 4b). COPD thus showed marked and hitherto unrecognized responsiveness to resistance training, contradicting previous suggestions of a negative impact of co-morbidities such as low cardiorespiratory fitness and chronic low-grade systemic inflammation.^8,38^

**Figure 4.**
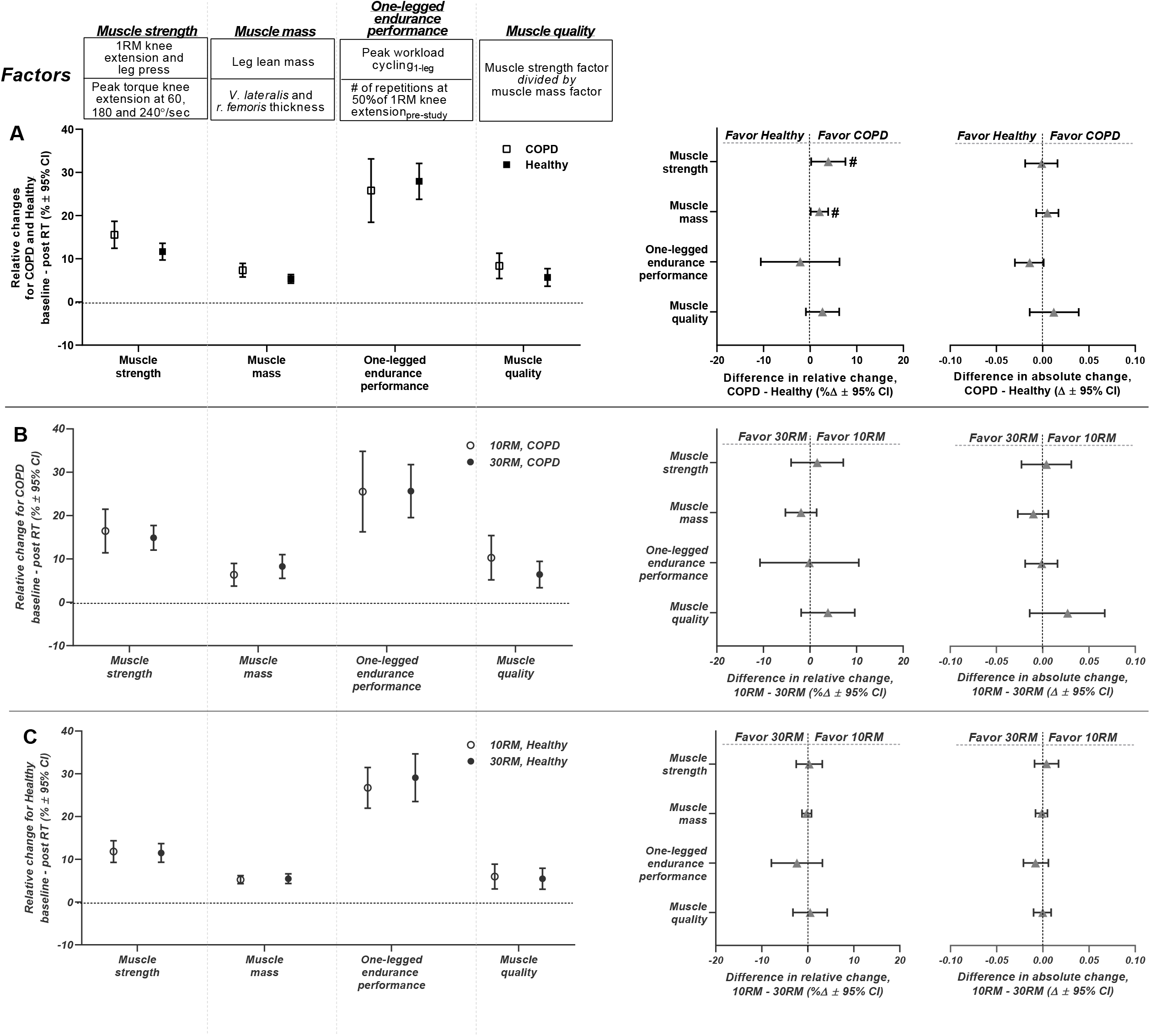
Effects of the resistance training intervention on lower-body muscle strength, lower-body muscle mass, one-legged endurance performance and lower-body muscle quality in COPD and Healthy. Each outcome domain is represented by a combined factor, computed from various performance assessments, as defined in the upper panel of the figure and previously described.^21^ (a) presents comparison of overall training effects between COPD and Healthy, measured as relative changes from baseline to after the resistance training intervention (per study cluster; left panel) and as relative and absolute differences in change scores between study clusters (right panels). In these analyses, high- and low-load resistance training (10RM and 30RM, respectively) were combined, warranted by the lack of differences between training load conditions in (b, c). COPD showed greater relative changes in muscle strength and muscle mass than Healthy. (b, c) presents comparison of effects of 10RM and 30RM resistance training in COPD (b) and Healthy (c) (i.e. per study cluster), measured as relative changes from baseline to after the intervention (left panels) and as relative and absolute differences in change scores between load conditions (right panels). #, statistically different effects of resistance training between COPD and Healthy. Data are presented as means with 95% confidence limits.

#### Cycling and functional performance

COPD and Healthy showed pronounced and similarly scaled training-associated improvements in whole-body endurance performance, measured as changes from baseline, including 6-min step test performance, 1-min sit-to-stand performance and maximal workload achieved during two-legged cycling (Figure 5). Surprisingly, COPD and Healthy also showed similar changes in performance for these outcome measures as absolute terms, with exception of 6-min step test performance (Δ-11 steps, Figure 5), for which Healthy showed larger improvements, arguably related to the considerable cardiorespiratory demand of this test, leaving COPD with morbidity-specific restraints. For other performance indices such as cycling economy and gross efficiency, which were measured using a one-legged cycling protocol, COPD showed larger relative improvements compared to Healthy (Δ4%, Figure 5). For these outcome measures, COPD, but not Healthy, displayed benefits of 10RM compared to 30RM training (Figure 5), corresponding to previously observed effects of heavy resistance training in healthy, young individuals.^39^

**Figure 5.**
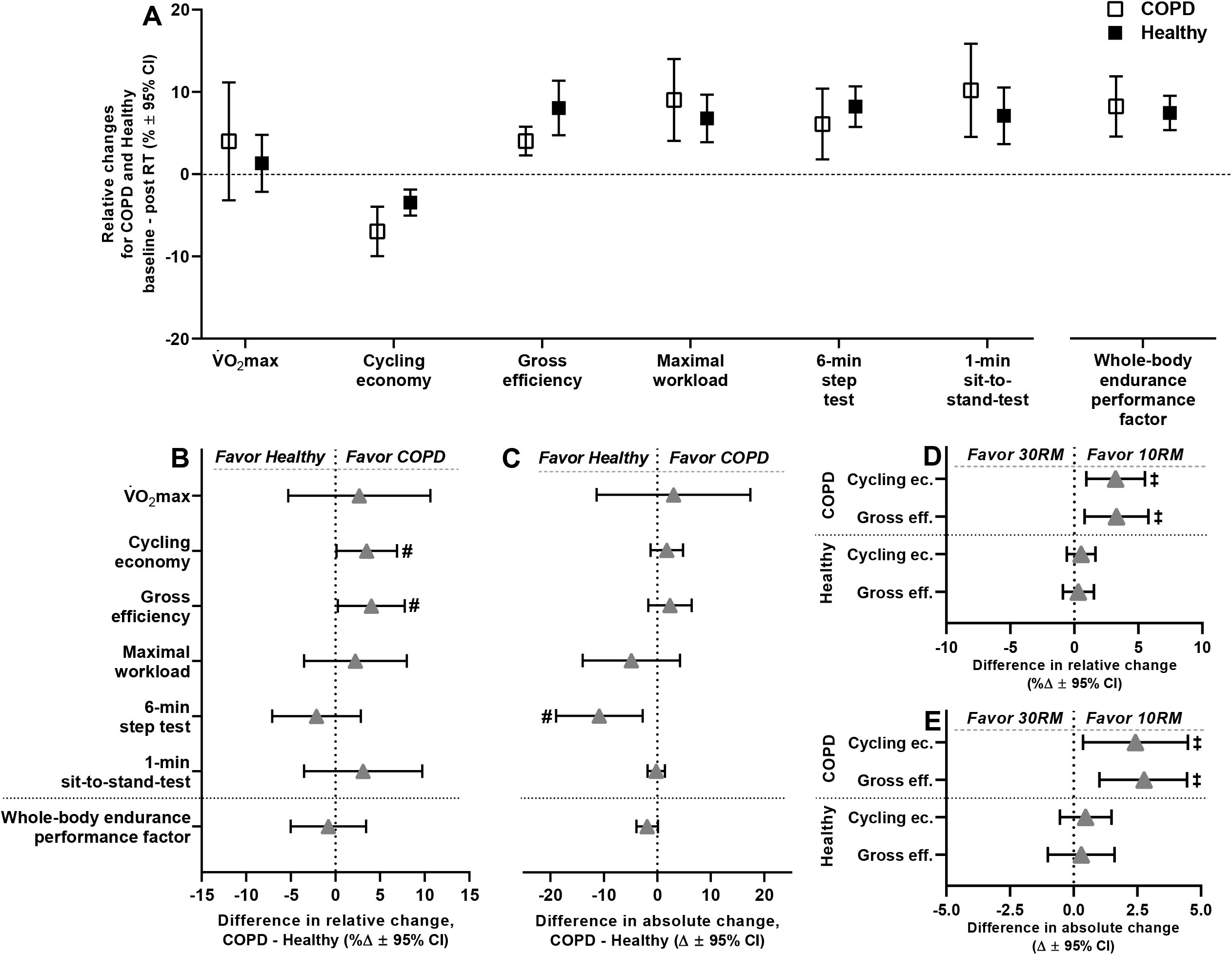
Comparison of the effects of the resistance training intervention on whole-body endurance performance in COPD and Healthy, presented as relative changes from baseline (per study cluster; a) and as relative and absolute differences in change scores between study clusters (b and c, respectively). Endurance measures included maximal oxygen consumption 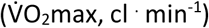and maximal workload (watts) achieved during two-legged cycling, cycling economy (cl. min^-1^) and gross efficiency measured during submaximal one-legged cycling, the number of steps achieved during 6-min step test, and the number of sit-to-stands achieved during a 1-min sit-to-stand test. COPD showed greater relative improvements in cycling economy and gross efficiency. For these outcome measures, COPD, but not Healthy, displayed benefits of high-load training (10RM) compared to low-load training (30RM) (d and e). Healthy showed greater absolute improvement in the number of steps achieved during the 6-min step test. COPD and Healthy showed similar relative and absolute training-associated changes in the whole-body endurance performance factor. #, statistically different response to resistance training between study clusters. ‡, statistically different response to 10RM and 30RM resistance training in study cluster. Data are presented as means with 95% confidence limits.

Together, these observations reiterate on the substantial benefits of resistance training for subjects with COPD, even for performance measures that pose large whole-body metabolic demands, which has previously been suggested to be irresponsive to such training.^40^ As such, it seems plausible that the observed improvements in 6-min step test performance, 1-min sit-to-stand performance and two-legged cycling were associated with improvements in work economy/gross efficiency and muscle strength, as neither COPD nor Healthy showed training-associated changes in maximal oxygen consumption (Figure 5), with improvements in anaerobic capacity being a potential contributor (not measured).

#### Muscle fiber characteristics

Whereas COPD and Healthy displayed similar increases in type II fiber CSA in *vastus lateralis* in response to resistance training (Δ-6%, p=0.438; Figure 6, upper panel), only Healthy showed increases in type I fiber CSA (16%), with no statistical difference being observed between study clusters. For Healthy, the increase in CSA was accompanied by increased myonuclei.fiber^-1^ in both fiber types (36%/25% for type I/II; Figure 7), leading to decreased myonuclear domain size estimates in type I fibers (−10%, Figure 7). For COPD, no such effects were observed (Figure 7). Despite the lack of difference between the two study clusters for these variables, the data hints at blunted plasticity of type I muscle fibers in COPD only, potentially relating to their altered biological characteristics at baseline or to blunted myonuclear accretion. Interestingly, in sub-analyses, the blunted type I responses in COPD seemed to be specific to 10RM training, with a tendency towards superior responses to 30RM training (Δ22%, p=0.060; Figure 6, middle panel). Such a phenomenon is supported by previous observations in responses to blood-flow-restricted low-load training,^41^ which arguably is mimicked by COPD subjects during low-load training, as they display inherent lowering of oxygen saturation in blood.

**Figure 6.**
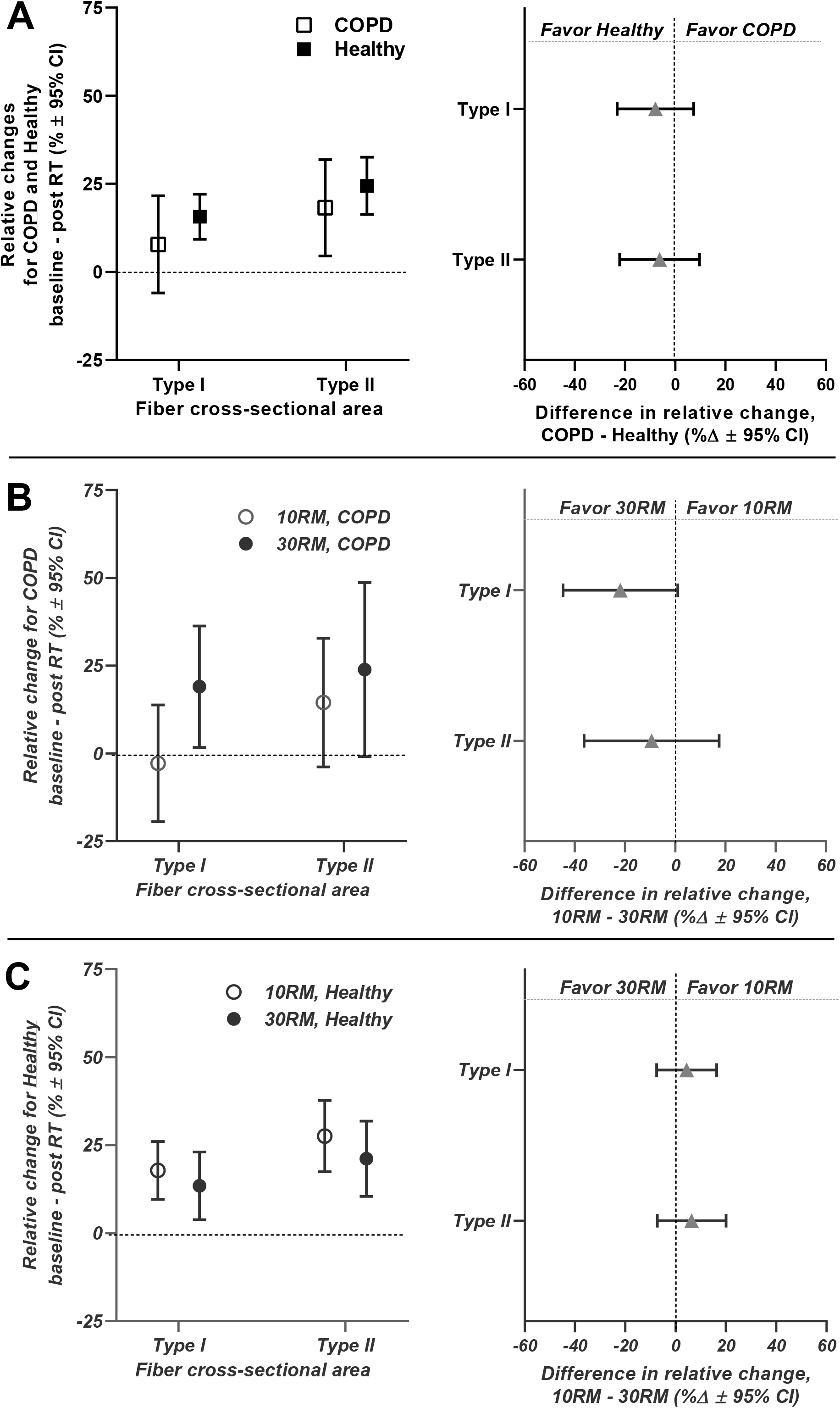
Effects of the resistance training intervention on cross-sectional area of muscle fiber types I and II in *m. vastus lateralis* in COPD and Healthy. (a) presents comparison of overall training effects on fiber CSA between COPD and Healthy, measured as relative changes from baseline to after the training intervention (per study cluster; left panel) and as relative differences in change scores between study clusters (right panel). In these analyses, high- and low-load resistance training (10RM and 30RM, respectively) were combined, warranted by the lack of significant differences between training load conditions in (b, c), though COPD tended to show higher efficacy of 30RM resistance training for changes in fiber type I CSA. (b, c) presents comparisons of effects of 10RM and 30RM resistance training on fiber CSA in COPD (b) and Healthy (c) (i.e. per study cluster), measured as relative changes from baseline to after the training intervention (left panels) and as relative and absolute differences in change scores between load conditions (right panels). Data are presented as means with 95% confidence limits.

**Figure 7.**
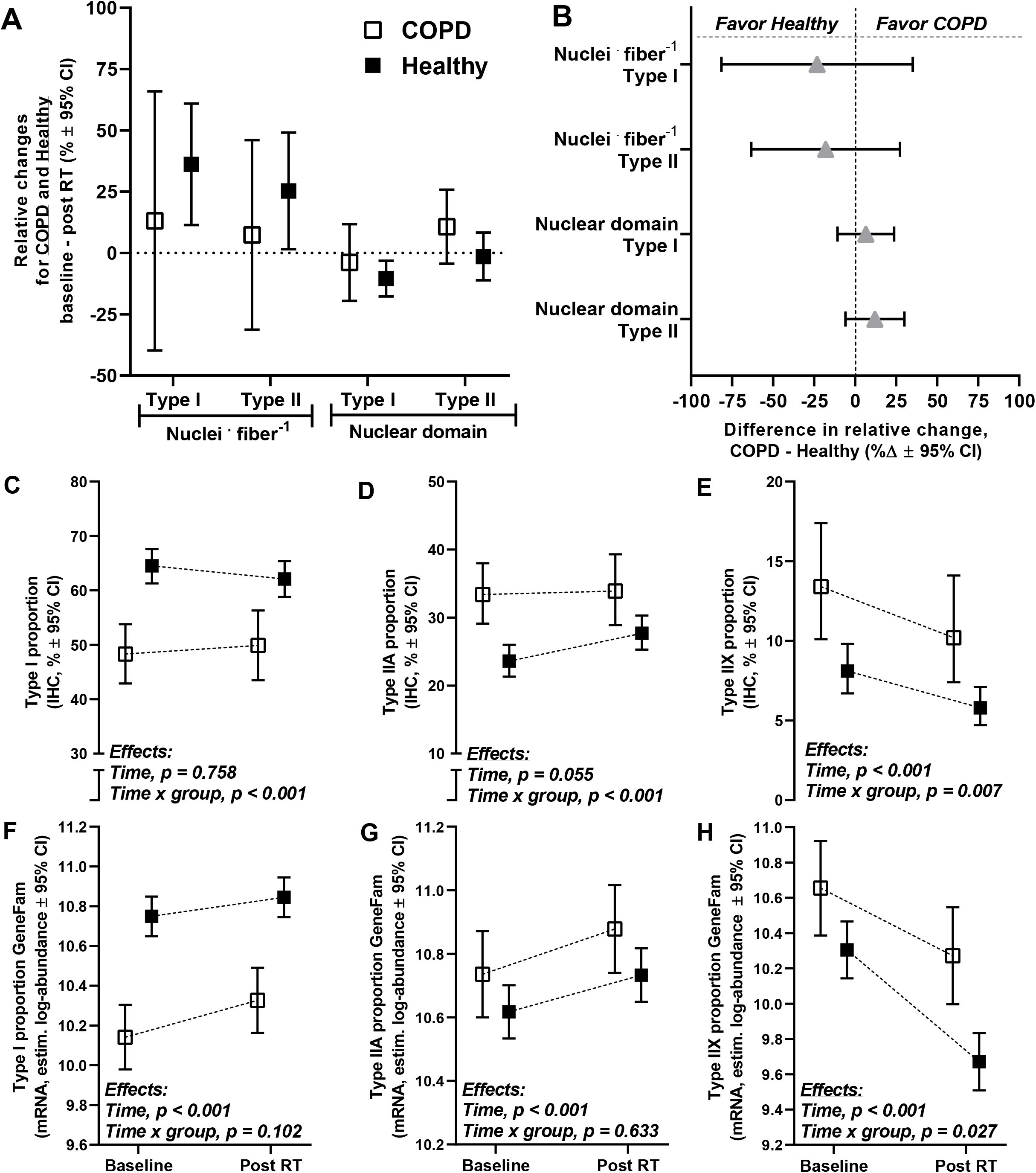
Comparisons of the effects of the resistance training intervention on changes in myonuclei *per* fiber and myonuclei domain in muscle fiber types I and II (a, b), and on changes in muscle fiber type proportions in COPD and Healthy, measured using immunohistochemistry (c-e) and qPCR (gene family profiling-normalized myosin heavy chain mRNA expression, f-h), as previously described.^24,49^ Myonuclei domain was calculated as mean fiber cross-sectional area divided by myonuclei *per* fiber. For myonuclei *per* fiber and myonuclei domain in muscle fiber types I and II, comparisons are presented as relative changes from baseline to after the training intervention (per study cluster; a) and as relative differences in change scores between study clusters (b). For muscle fiber type proportions, data are presented as adjusted values at baseline and after the training intervention (Post RT), and results are presented as the effect of the training intervention for the study clusters combined and its interaction with study clusters (c-h). For myonuclei variables, no training-associated differences were observed between study clusters. Both COPD and Healthy displayed training-associated reductions in proportions of type IIX muscle fibers, measured using both immunohistochemistry and qPCR. Intriguingly, while this reduction was greater in COPD when measured at the protein level (immunohistochemistry), it was greater in Healthy when measured at the mRNA level (qPCR), indicating differentially regulated muscle fiber shifting in COPD and Healthy. Data are presented as means with 95% confidence limits.

Both COPD and Healthy displayed training-associated reductions in type IIX muscle fiber proportions (Figure 7). While this reduction was more pronounced in COPD when measured at the protein level (immunohistochemistry), it was more pronounced in Healthy when measured at the mRNA level, suggesting differential orchestration of muscle fiber shifts between study clusters, possibly relating to their inherently different muscle fiber proportions at baseline.

#### Muscle RNA content

In general, COPD and Healthy showed similar increases in ribosomal RNA abundance *per* unit muscle tissue weight, measured as both total RNA and rRNA expression, and measured after both 3½ week (1.19/1.29 and 1.15/1.16 fold increases, total RNA/rRNA abundances) and after finalization of the training intervention (1.13/1.18 and 1.05/1.17 fold increases) (Figure 8). While these changes in ribosomal RNA content were generally similar between COPD and Healthy, a few noteworthy differences were evident, including a more robust early increase in 45s pre-rRNA abundance in COPD (Figure 8) and a trend towards reduced changes in response to 13 weeks training in COPD, which led to the absence of time effects for all rRNA species. The early increases in ribosomal content seen in both COPD and Healthy resemble those typically seen after similar interventions in untrained young individuals,^24^ and may be important for muscle growth capabilities over the entirety of the study period,^24,42^ accommodating increases in protein synthesis capacity, thus potentially contributing to the pronounced muscular responses to resistance training seen in both study clusters.

**Figure 8.**
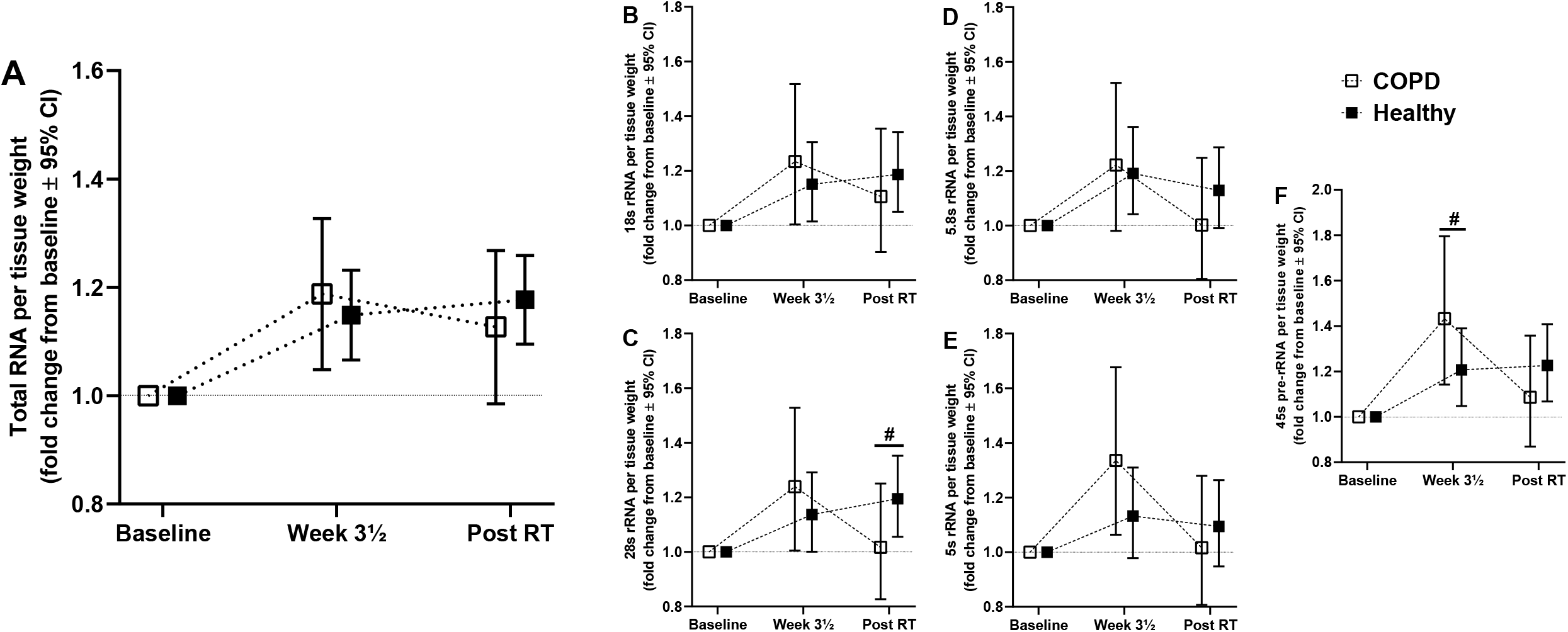
Effects of the resistance training intervention on total RNA content (a) and rRNA expression (b-f) in *m. vastus lateralis* of COPD and Healthy. Data are presented as fold changes from baseline to Week 3½ (Post-intro RT; seven training sessions) and to after the training intervention (Post RT; 26 training sessions). Total RNA (a), 18s rRNA (b), 28s rRNA (c), 5.8s rRNA (d) 5s rRNA (e) and 45s pre-rRNA (f) abundances. Total RNA- and qPCR-analyses were assessed as per-amounts of tissue weight, as previously described.^21,24^ *#*, statistical difference in fold change between COPD and Healthy (alpha level, *p*<0.05). Data are presented as means with 95% confidence limits.

In both COPD and Healthy, resistance training led to marked changes in mRNA transcriptome profiles, with 499 and 312 differentially expressed genes being observed after 3½ and 13 weeks of resistance training, respectively (for general information about transcriptomic responses, see Mølmen *et al*.^21^). Overall, at the single-gene level, no transcripts showed differential responses to training between the two study clusters, neither at 3½ weeks nor at 13 weeks, despite clear differences in transcriptome profiles at baseline (Figure 3a, online data supplement, Table E3). In contrast, enrichment analyses revealed traces of differential changes (Figure 3c, Table 2 and online data supplement, Table E4), with COPD showing more pronounces increases in expression of genes relating to *oxidative phosphorylation* after 3½ weeks (GSEA), and, in particular, more pronounced decreases in genes associated with *myogenesis* after 13 weeks (consensus) (Figure 3c, Table 2). Interestingly, as these two gene sets represented the most prominent differences between COPD and Healthy at baseline (Figure 3a-b), and as resistance training led to directional changes that mitigated these differences, training arguably shifted the COPD phenotype in a healthy direction.

#### Blood and health-related outcomes

Overall, COPD and Healthy showed similar training-associated increases in whole-body and appendicular lean mass (Table 3). This was accompanied by increased appendicular skeletal muscle mass index relative to the sex-specific mean of young, healthy adults (COPD, from 84% to 86%; Healthy, from 95% to 97%), suggesting that the intervention was effective for reversing age-related decline in muscle mass. For blood variables such as markers of systemic inflammation and hormone, lipid and iron biology, no noteworthy effects were observed of the intervention, nor were any differential changes observed between COPD and Healthy (Table 3).

#### Lung function

For COPD, the training intervention did not affect any of the lung function variables (Table 4), implying no effects on this core epidemiological trait. This seems reasonable given the irreversible nature of the respiratory impairments of COPD, contradicting the beneficial effects observed in Hoff *et al*.^14^ In contrast, for Healthy, the intervention was associated with reduced FVC and FEV_1_ (−2.7% and −1.5%, respectively). Rather than being a consequence of the intervention protocol *per se*, this may be due to a general age-related decline, as the magnitude of the changes resemble those seen in corresponding age cohorts over a similar time frame.^43^

**Table 4.** Effects of the training intervention on lung function in COPD and Healthy, assessed as changes from baseline to after completion of the study (per study cluster) and as differential changes between study clusters.

| | COPD | | Time effect<br>p<0.05) | Healthy | | Time effect<br>(p<0.05) | $\Delta$ COPD vs $\Delta$ healthy<br>(p-value) |
| --- | --- | --- | --- | --- | --- | --- | --- |
|  | Baseline | Post RT |  | Baseline | Post RT |  |  |
| FVC (L) | 3.3 ± 0.9 | 3.2 ± 0.9 | No | 3.6 ± 0.9 | 3.5 ± 0.8 | Yes ↓ | 0.189 |
| FEV<sub>1</sub> (L · sec<sup>-1</sup>) | 1.5 ± 0.4 | 1.5 ± 0.4 | No | 2.7 ± 0.7 | 2.7 ± 0.6 | Yes ↓ | 0.243 |
| FEV<sub>1</sub> (% predicted) | 56 ± 11 | 58 ± 13 | No | 103 ± 16 | 103 ± 16 | No | 0.138 |
| FEV<sub>1</sub>/FVC (%) | 47 ± 8 | 48 ± 10 | No | 75 ± 6 | 76 ± 6 | No | 0.714 |
| PEF (L · sec<sup>-1</sup>) | 5.0 ± 1.6 | 5.1 ± 1.6 | No | 7.8 ± 2.1 | 7.6 ± 2.2 | No | 0.238 |

FVC, forced vital capacity; FEV<sub>1</sub>, forced expiratory volume in one second; PEF, peak expiratory flow; $\Delta$ , change score. Alpha level at $p < 0.05$ . Values are means with standard deviation.

#### Health-related quality of life

For COPD, the intervention was associated with marked improvements in several aspects of health-related quality of life (Table 5). These included reduced experience of limitations of physical functioning and improved social function and mental health, with only marginal effects being seen in Healthy. While these changes of course may be directly related to the resistance training intervention, they may also be related to other aspects of the study protocol, such as performing training sessions in a social setting and the close follow-up each participant received from study personnel. As the intervention was conducted without a control group (not receiving the intervention protocol), caution is warranted for interpretation of these data.

**Table 5.** Effects of the training intervention on health-related quality of life in COPD and Healthy, measured using COPD Assessment Test (CAT; COPD-only) and the 36-item Short Form Health Survey (SF-36; all participants), and assessed as changes from baseline to after completion of the study (per study cluster; CAT and SF-36) and as differential changes between study clusters (SF-36).

| | <u>COPD</u> | | | <u>Healthy</u> | | | $\Delta$ COPD vs<br>$\Delta$ Healthy<br>(*P* value) |
| --- | --- | --- | --- | --- | --- | --- | --- |
|  | Baseline | Post RT | Time effect<br>*P*<0.05) | Baseline | Post RT | Time effect<br>(*P*<0.05) |  |
| **COPD assessment Test™ score (0-40)** | 16.6 ± 6.8 | 16.4 ± 6.8 | No | - | - | - | - |
| **Short Form (36) Health Survey (0-100)** |  |  |  |  |  |  |  |
| Physical function * | 63 ± 19 | 67 ± 18 | No | 90 ± 14 | 92 ± 12 | No | 0.321 |
| Role physical * | 43 ± 34 | 59 ± 37 | Yes ↑ | 87 ± 25 | 94 ± 18 | No | 0.226 |
| Bodily pain | 71 ± 27 | 82 ± 19 | Yes ↑ | 79 ± 21 | 80 ± 19 | No | 0.070 |
| General health * | 48 ± 20 | 56 ± 19 | No | 75 ± 18 | 80 ± 12 | No | 0.208 |
| Vitality * | 52 ± 16 | 57 ± 13 | No | 72 ± 18 | 78 ± 11 | Yes ↑ | 0.509 |
| Social function * | 74 ± 23 | 84 ± 16 | Yes ↑ | 90 ± 18 | 94 ± 13 | No | 0.280 |
| Role emotional * | 65 ± 39 | 84 ± 26 | Yes ↑ | 93 ± 19 | 96 ± 15 | No | 0.059 |
| Mental health * | 77 ± 13 | 84 ± 13 | Yes ↑ | 86 ± 11 | 89 ± 8 | Yes ↑ | 0.196 |

\*, difference between COPD and Healthy at baseline; ↑, significant increase from baseline to after the training intervention (Post RT). Alpha level at *p*<0.05. Values are means with standard deviation.

## Concluding remarks

COPD-related pathophysiologies, such as reduced testosterone,^3^ vitamin D^4^ and oxygen saturation levels^6,7^ in blood and elevated levels of low-grade inflammation,^8^ are generally believed to drive metabolism into a chronic catabolic state.^3,6,9^ This has been suggested to lead to impaired responses to lifestyle interventions such as resistance training,^6,44^ which are essential measures for preventing and treating disease-related reductions in skeletal muscle mass and strength, counteracting escalation into serious conditions such as pulmonary cachexia.^17^ Despite this general belief, the presence of impaired training responsiveness in COPD is not backed by experimental data, and there is no *de facto* evidence for such limited responses to exercise training. To date, a mere single study has compared responses between COPD and healthy control subjects,^18–20^ and as such failing to lend support to the prevailing view. In the present study, we largely disavow the myth of impaired responsiveness to training in COPD, measured as responses to a 13-week whole-body resistance training intervention, conducted using an exhaustive follow-up and testing protocol, including extensive test-retest validations (see Mølmen *et al*.^21^). Whereas COPD participants displayed clear and well-known disease-related aberrancies compared to Healthy at baseline, including altered skeletal muscle characteristics and elevated levels of systemic inflammation, they showed similar or superior improvements for virtually every measure of health, performance and biology. Specifically, COPD showed greater relative improvements in core outcome domains such as lower-body muscle strength and mass, and similar relative improvements in muscle quality, one-legged endurance performance and whole-body endurance performance. These similarities between COPD and Healthy were also evident as absolute change terms, suggesting that the improvements seen in COPD was decoupled from the compromised levels at baseline. These observations were accompanied by similar alterations in muscle biology, including changes in hallmark traits such as muscle fiber characteristics, rRNA content and transcriptome profiles. Together, these data suggest that the COPD etiology does not lead to impaired responsiveness to resistance training, at least not for skeletal muscle characteristics, and at least not in the enrolled cluster of COPD participants (GOLD grade II-III) or within the time frame of the study.

During planning of the study protocol, two strategies were implemented to resolve the hypothesized, albeit now rejected, negative impact of COPD-specific pathophysiologies for the efficacy of resistance training. *First*, as vitamin D insufficiency is common among COPD subjects,^4^ and has been suggested to contribute to development of the postulated anabolic resistance,^45^ dietary habits were manipulated to investigate the effects of vitamin D_3_ supplementation. Contrary to our hypothesis, vitamin D_3_ did not enhance responses to resistance training for any of the outcome variables.^21^

*Second*, the resistance training protocol was conducted using two different training modalities, 10RM and 30RM resistance training, performed in a contralateral manner. The efficacies of these training modalities were initially hypothesized to be dissimilarly affected by COPD-related pathophysiologies, as they convey muscular adaptations through different signaling cues in the cellular environment (i.e. mechanical tension vs metabolic perturbation),^46^ and may thus well be differentially affected by extracellular signaling such as inflammation and oxygen availability. While this hypothesis was rejected for all core outcome domains, with no differences being observed between the two training modalities and no evidence being found to support the presence of impaired training responsiveness, a noteworthy observation was made for muscle fiber-specific traits. Specifically, in COPD, 10RM training was associated with blunted growth of type I muscle fiber CSA, a phenomenon that was not observed for responses to 30RM training, suggesting that 30RM offers benefits for muscle fiber type I hypertrophy. Apart from this, 10RM was observed to lead to greater improvements in cycling economy and gross efficiency in COPD. These arguably contradicting observations warrant further study.

In conclusion, COPD performing a well-tolerated 13-week resistance training program displayed pronounced improvements for a range of health and muscle functional and biological variables, resembling or exceeding those seen in Healthy. Hence, COPD did not lead to impaired responsiveness to exercise training, which rather posed a potent measure to relieve COPD-related co-morbidities.

Acknowledgments

The authors would like to express their gratitude to all students involved in the study for invaluable assistance during intervention follow-up and data sampling. The authors also acknowledge the contributions to the study from MD Bjørn S. Svensgaard (Innlandet Hospital Trust), conducting the pre-inclusion consultations for the participants with COPD, biomedical laboratory technician Randi Sivesind (Innlandet Hospital Trust), performing the blood analyzes, Prof. Olivier Seynnes (Norwegian School of Sport Sciences) for instructions and education in ultrasound assessments, and Thomas Urianstad, Peter Nore Bengtsson, Gudmund Storlien, Joar Hansen and Anne Sofie Lofthus for valuable support. Finally, yet importantly, we would like to thank all study participants for their effortful and dedicated contributions.

## Authors’ contributions

KSM and SE developed the project, with input from GSF, TJR, BRR and TR. KSM led the study intervention, including coordination and conduction of exercise training and testing, with aid from DH, GSF, BRR and SE. MG and TJR planned, organized and conducted participant recruitment and performed medical screening. BM and RL planned, organized and conducted lung spirometry and DXA measurements. KSM, DH, LK, MH, YK, RA and SE planned and performed muscle biological analyses. KSM, DH and SE planned and performed data analyses, with input from YK and RA. KSM, TR and SE drafted the manuscript. All authors provided useful input to data interpretation and contributed to drafting and finalizing the manuscript. All authors have read and approved the final manuscript.

## Funding information

The study was funded by Inland Norway University of Applied Sciences, Innlandet Hospital Trust (grant number 150339, SE) and Regional Research Fund Inland Norway (grant number 298419, SE).

## At a Glance Commentary

### Scientific Knowledge on the Subject

Subjects with chronic obstructive lung disease (COPD) are prone to accelerated decay of muscle strength and mass with advancing age. This is mediated by systemic pathophysiologies, such as elevated levels of low-grade inflammation, and reduced levels of testosterone, vitamin D and oxygen saturation levels in blood. This has been suggested to hinder responses to lifestyle interventions such as resistance training, though this notion is largely unstudied.

### What this Study Adds to The Field

In the present study, we investigated the presence of impaired training responsiveness in COPD, measured as responses to a 13-week whole-body resistance training intervention compared to healthy participants. The study was conducted using an exhaustive follow-up and testing protocol, including extensive test-retest validations. Contrary to our hypothesis, COPD participants displayed similar or superior improvements in virtually every measure of health, performance and biology compared to healthy participants, measured as both relative and absolute change terms. Based on these data, it seems prudent to reject the notion that COPD is associated with impaired responsiveness to resistance exercise training, and rather encourage resistance training as a potent measure to relieve COPD-related co-morbidities.

## Data Availability

Data available on request from the authors.

## Notes

### Competing Interest Statement

The authors have declared no competing interest.

### Clinical Trial

NCT02598830

### Author Declarations

This study was approved by the Regional Committee for Medical and Health Research Ethics - South-East Norway (reference nr: 2013/1094)

