## Supplementary material for "Chronic Obstructive Pulmonary Disease Does Not Impair Responses to Resistance Training": Online Data Supplement

***Data analyses and statistics***

For continuous variables, linear mixed-effects models were used to examine differences between COPD and Healthy, both at baseline and as responses to resistance training. For the latter, relative and absolute changes from baseline were defined as dependent variables, with COPD/Healthy being defined as the fixed effect. Analyses included evaluation of interaction effects with training load (repeated measures/observations from the high- and low-load training leg were added to the model for unilateral outcome measures) and sex. The effects of sex were implemented into the models. Time effects were examined using mixed modelling, with the dependent variable and time points being defined as repeated measures/observations.

For non-continuous variables (fiber type proportions, rRNA/mRNA content), generalized linear mixed-effects models were used. In transcriptome analyses, genes were regarded as differentially expressed when the absolute log2 fold-change/difference were greater than 0.5 and the adjusted *p*-value (false discovery rate adjusted *per* model coefficient) was below 5%.[E1] Moreover, enrichment analyses were performed on hallmark, KEGG and gene ontology gene sets, using two approaches. First, a non-parametric rank test was performed based on gene-specific minimum significant differences (MSD). Second, gene set enrichment analysis (GSEA) was performed to quantify directional regulation of the gene set. Consensus results were interpreted as having larger biological meaning, while Hallmark was providing the most meaningful stand-alone interpretation, as it reduces the analytical noise by taking into account genes that overlap between gene sets.[E2] All gene sets were retrieved using the molecular signature database (version 7.1.).[E3] Overview of gene enrichment analyses with exact *p*-values are presented in e-Table 4.

For all immunohistochemical variables, statistical models were weighted for numbers of counted fibers *per* biopsy.

To achieve reliable assessment of core outcome domains, and thus to lower the risk of statistical errors, combined factors were calculated for outcome measures relating to lower-body muscle strength, lower-body muscle mass, one-legged endurance performance and whole-body endurance performance, as previously described.[E4] For more information, see e-Table 1.

Statistical significance was set to $p<0.05$. In both text and figures, data are presented as adjusted, marginal means, with or without 95% confidence intervals, unless otherwise stated. Statistical analyses were performed using SPSS Statistics package version 24 (IBM, Chicago, IL, USA) and R software.[E5] Figures were made using Prism Software (GraphPad 8, San Diego, CA, USA) and R software.[E5]

***The quality and general efficacy of the resistance training protocol***
The training intervention was associated with low drop-out rates (n=4, ~5%; COPD, n=2) and high adherence to the protocol (COPD, 97%; Healthy, 98%), likely ensured by close follow-up from qualified personnel during all training sessions. Training volume increased steadily throughout the intervention (Figure 2), with Healthy showing larger increases than COPD from week 1 to Week 4 (COPD, 18%; Healthy, 28%; p=0.023), but not from Week 1 to Week 8 (COPD, 42%; Healthy, 54%; p=0.091) or from Week 1 to Week 13 (COPD, 56%; Healthy, 70%; p=0.142) (Figure 2). In both COPD and Healthy, training was associated with progressive increases in perceived exercise intensities using the Borg RPE-scale [E6] (Figure 2), with no difference between study groups. Healthy-only showed increase in relative training load (% of one repetition maximum, 1RM) during the intervention (Figure 2).

To ensure valid assessment of the efficacy of the training protocol for muscle-related characteristics, two precautionary measures were made, as previously described.[E4] First, for muscle strength and performance, baseline levels were defined as values collected after 3½ weeks of introduction to resistance training (Figure 2). At this time point, the initial adaptations to training were likely to have occurred, preferably non-hypertrophic adaptations relating to technical, psychological and neural learning effects.[E7] To further minimize the confounding effects of these variables, all participants conducted a series of repeated tests prior to baseline tests (five repeated 1RM and muscular performance tests in knee extension and chest press conducted prior to introduction to resistance training; three repeated tests for 1RM leg press; e-Figure 1). For 1RM knee extension and chest press (but not for 1RM leg press and muscular endurance tests), these learning effects were greater in COPD: 14% ± 20% in knee extension (vs 5% ± 12% in Healthy, p=0.006) and 10% ± 11% in chest press (vs 3% ± 9% in Healthy, p=0.004). For 1RM chest press, this difference between study groups was present in analyses of absolute change scores (p=0.010), but not for knee extension (p=0.056). Even after adopting these strict criteria for baseline assessment of muscle strength, the general efficacy of the resistance training intervention was somewhat higher than expected based on previous studies in both study groups (data from the two legs combined): e.g. 1RM knee extension (0.9% $\cdot$ session$^{-1}$/0.8% $\cdot$ session$^{-1}$, COPD/Healthy) and 1RM leg press (1.4% $\cdot$ session$^{-1}$/1.3% $\cdot$ session$^{-1}$, COPD/Healthy).[E8–10] Second, for the core outcome domains of the study (muscle strength, muscle mass, one-legged endurance performance and whole-body endurance performance), end-point analyses were based on combined factors computed from the entire range of singular assessments rather than on any singular outcome measure, as previously recommended[E11] (for in-depth information, see e-Table 1). This was done to optimize the reliability of our assessment and thus to lower the risk of statistical errors.[E4]

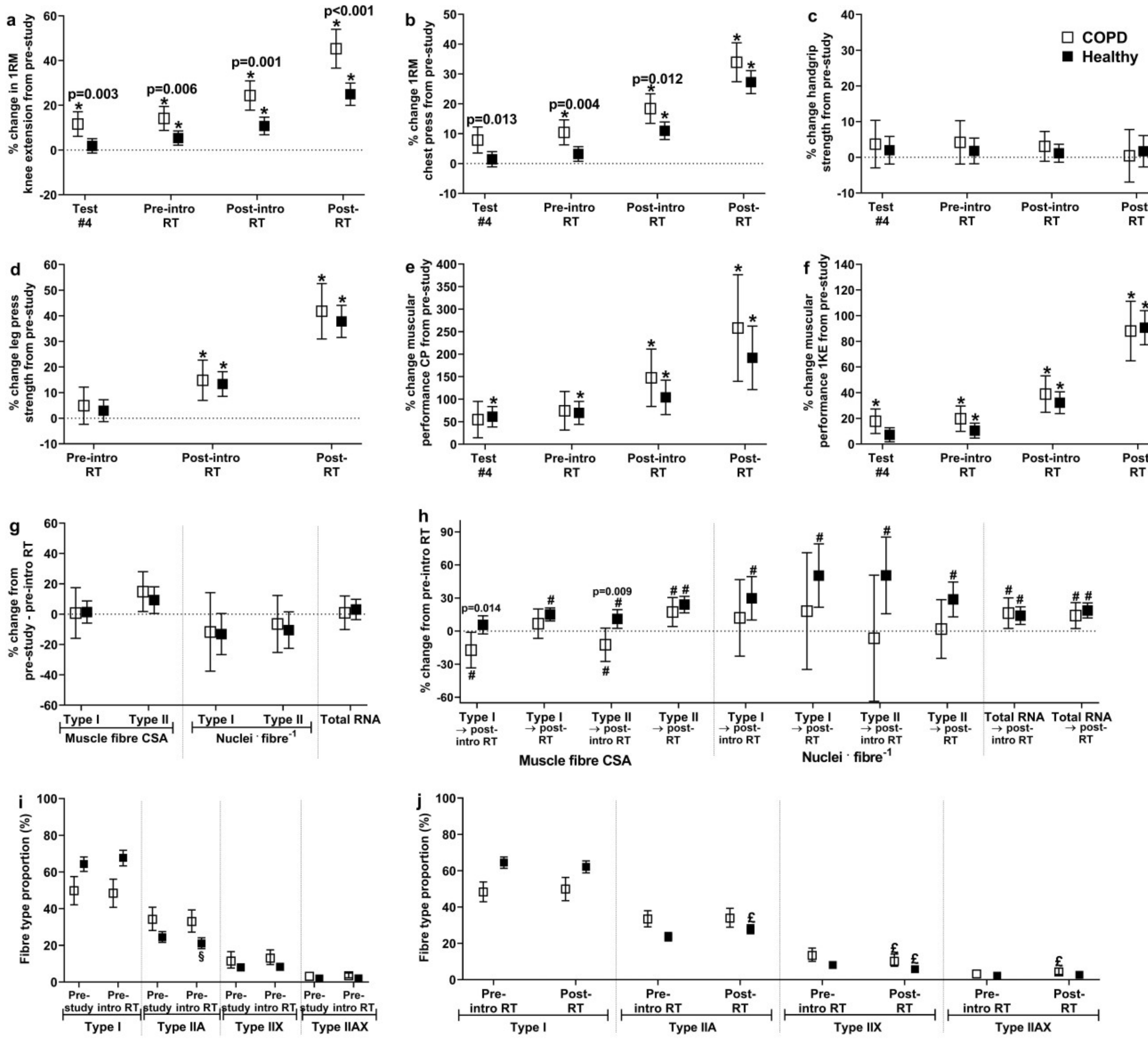

**Figure E1.** General efficacy of the study intervention measured as changes in one repetition maximum one-legged knee extension (a), one repetition maximum chest press (b), grip strength (c), one repetition maximum one-legged leg press (d), muscular performance chest press (number of repetitions at 50% of pre-study one repetition maximum; e), muscular performance one-legged knee extension (number of repetitions at 50% of pre-study one repetition maximum; f), muscle fiber cross-sectional area (CSA; type I and type II fibers), nuclei per fiber and total RNA in *m. vastus lateralis* (from pre-study to baseline/pre-introduction resistance training, dominant leg only, g; from baseline/pre-introduction resistance training to post-introduction resistance training and post resistance training (i.e. after study intervention), h), and muscle fiber type proportions (immunohistochemistry; pre-study and pre-introduction resistance training/baseline, dominant leg only, i; pre-introduction resistance training/baseline and post resistance training, both legs, j). Test 4, test performed in Week 8 (see Figure 2); RT, resistance training; *, significant change from pre-study; #, significant change from baseline/pre-introduction resistance training; £, significant different from baseline/pre-introduction resistance training. Significant differences between study clusters are marked with *p*-values.

**Table E1.** Computed factors for the core outcome domains lower-body muscle mass, lower-body muscle strength, one-legged endurance performance and whole-body endurance performance. Each factor consists of multiple singular outcome measures. First, for each outcome measure, each subject's value (pre and post) was normalized to the highest recorded value during the study conduct, thus providing values <1. Then, for each subject, an ultimate factor was computed for lower-body muscle mass, lower-body muscle strength, one-legged endurance performance and whole-body endurance performance, respectively, calculated as the mean of normalized values for the various variables included.

| ***Lower-body muscle mass factor*** | | | | | | | | | | |
|---|---|---|---|---|---|---|---|---|---|---|
| | *Included variables* | *Explanation* | *Baseline (avg ± SD)* | *Post intervention (avg ± SD)* | *Estimate, change (95% CI)* | *Main effect of time (p value)* | *Correlation with factor at baseline, r value (p value)* | *Correlation for change score with change score for factor, r value (p value)* | *Eigenvalue* | *% variance explained* |
| | 1. Muscle thickness | The combined measure of muscle thickness of *vastus lateralis* and *rectus femoris* | 0.60 (0.10) | 0.66 (0.11) | 0.06 (0.05, 0.07) | <0.001 | 0.83 (<0.001) | 0.84 (<0.001) | - | - |
| | 2. Leg lean mass | Lean mass in the legs | 0.64 (0.15) | 0.65 (0.15) | 0.01 (0.01, 0.02) | <0.001 | 0.92 (<0.001) | 0.63 (<0.001) | - | - |
| | **Lower-body muscle mass factor** | - | **0.63 (0.11)** | **0.66 (0.12)** | **0.04 (0.03, 0.04)** | **<0.001** | - | - | **1.10** | **55** |
| ***Lower-body muscle strength factor*** | | | | | | | | | | |
| | *Included variables* | *Explanation* | *Baseline (avg ± SD)* | *Post intervention (avg ± SD)* | *Estimate, change (95% CI)* | *Main effect of time (p value)* | *Correlation with factor at baseline, r value (p value)* | *Correlation for change score with change score for factor, r value (p value)* | *Eigenvalue* | *% variance explained* |
| | 1. Leg muscle strength | The combined measure of 1RM knee extension and leg press | 0.44 (0.14) | 0.52 (0.15) | 0.08 (0.07, 0.09) | <0.001 | 0.95 (<0.001) | 0.78 (<0.001) | - | - |
| | 2. Leg muscle torque | The combined measure of torque (Nm) achieved during knee extension at 60°, 180° and 240°/sec | 0.48 (0.16) | 0.51 (0.17) | 0.03 (0.02, 0.04) | <0.001 | 0.97 (<0.001) | 0.73 (<0.001) | - | - |
| | **Lower-body muscle strength factor** | - | **0.46 (0.14)** | **0.52 (0.15)** | **0.06 (0.05, 0.06)** | **<0.001** | - | - | **1.13** | **57** |
| ***One-legged endurance performance factor*** | | | | | | | | | | |
| | *Included variables* | *Explanation* | *Baseline (avg ± SD)* | *Post intervention (avg ± SD)* | *Estimate, change (95% CI)* | *Main effect of time (p value)* | *Correlation with factor at baseline, r value (p value)* | *Correlation for change score with change score for factor, r value (p value)* | *Eigenvalue* | *% variance explained* |
| | 1. Muscle performance | Number of repetitions at 50% of 1RM knee extension | 0.13 (0.03) | 0.22 (0.09) | 0.09 (0.08, 0.10) | <0.001 | 0.60 (<0.001) | 0.92 (<0.001) | | |
| | 2. Maximal power output | Maximal power output achieved during one-legged cycling | 0.49 (0.17) | 0.52 (0.17) | 0.04 (0.03, 0.04) | <0.001 | 0.99 (<0.001) | 0.29 (<0.001) | | |
| | **One-legged endurance performance factor** | - | **0.31 (0.09)** | **0.37 (0.11)** | **0.06 (0.06, 0.07)** | **<0.001** | | | **1.11** | **56** |

| | | | | | | | | | | |
|---|---|---|---|---|---|---|---|---|---|---|
| ***Whole-body endurance performance factor*** | | | | | | | | | | |
| | ***Included variables*** | ***Explanation*** | ***Baseline (avg ± SD)*** | ***Post intervention (avg ± SD)*** | ***Estimate, change (95% CI)*** | ***Main effect of time (p value)*** | ***Correlation with factor at baseline, r value (p value)*** | ***Correlation for change score with change score for factor, r value (p value)*** | ***Eigenvalue*** | ***% variance explained*** |
| | Maximal power output | Maximal power output achieved during bicycling | 0.44 (0.17) | 0.47 (0.18) | 0.03 (0.02, 0.04) | <0.001 | 0.87 (<0.001) | 0.54 (<0.001) | - | - |
| | 6-min step test | Number of steps achieved during a 6-min test | 0.59 (0.17) | 0.63 (0.19) | 0.04 (0.03, 0.06) | <0.001 | 0.96 (<0.001) | 0.64 (<0.001) | - | - |
| | 1-min sit-to-stand test | Number of sit-to-stands achieved during a 1-min test | 0.58 (0.13) | 0.62 (0.14) | 0.04 (0.03, 0.06) | <0.001 | 0.86 (<0.001) | 0.76 (<0.001) | - | - |
| | **Whole-body endurance performance factor** | - | 0.54 (0.14) | 0.58 (0.15) | 0.04 (0.03, 0.05) | **<0.001** | - | - | **1.29** | **43** |

**Table E2.** Comparison of baseline characteristics between COPD and Healthy

| Variable | COPD | Healthy | Sex-adjusted estimated differences between groups: COPD – Healthy (95 % CI) | P-value |
|---|---|---|---|---|
| *Maximal oxygen consumption* | | | | |
| Two-legged cycling $VO_2$max, ml · min$^{-1}$ | 1478 ± 473 | 2370 ± 682 | -1193 (-930, -1456) | < 0.001 # |
| Two-legged cycling $VO_2$max, ml · kg$^{-1}$ · min$^{-1}$ | 19.0 ± 5.7 | 30.9 ± 7.3 | -13.6 (-9.5, -17.7) | < 0.001 # |
| One-legged cycling $VO_2$max, ml · min$^{-1}$ | 1221 ± 304 | 1875 ± 480 | -779 (-638, -921) | < 0.001 # |
| One-legged $VO_2$max/two-legged $VO_2$max (%) | 92 ± 12 | 81 ± 9 | 13 (20, 6) | < 0.001 # |
| *Body composition* | | | | |
| Lean mass, kg | 46.7 ± 9.9 | 48.1 ± 10.0 | -6.4 (-3.9, -8.9) | < 0.001 # |
| Appendicular skeletal muscle mass (ASMM), kg | 20.3 ± 5.3 | 21.6 ± 5.0 | -3.8 (-2.4, -5.2) | < 0.001 # |
| ASMM index, appendicular skeletal muscle mass (kg)/m$^2$ | 6.87 ± 1.30 | 7.42 ± 1.12 | -1.1 (-0.6, -1.5) | < 0.001 # |
| ASMM index, % of sex-specific means of young adults | 84.2 ± 12.7 | 95.4 ± 10.1 | -13 (-7, -19) | < 0.001 # |
| Sarcopenia, no., ASMM index >2 SD below the sex-specific means of young adults £ | 9/20 | 2/58 | - | - |
| Fat mass, kg | 26.4 ± 11.7 | 25.3 ± 9.3 | 1.1 (6.8, -4.6) | 0.703 |
| Visceral fat, kg | 1.6 ± 1.2 | 1.1 ± 0.9 | 0.2 (0.7, -0.3) | 0.412 |
| Bone mineral density (g · cm$^2$) | 1.13 ± 0.21 | 1.15 ± 0.16 | -0.09 (-0.03, -0.16) | 0.007 # |
| *Whole-body performance/functional measures* | | | | |
| Peak work rate, W | 103 ± 41 | 198 ± 57 | -113 (-92, -134) | < 0.001 # |
| Six-minute-step-test, steps | 120 ± 38 | 200 ± 39 | -83 (-61, -104) | < 0.001 # |
| One-minute sit-to-stand-test | 21 ± 5 | 30 ± 5 | -9 (-6, -12) | < 0.001 # |
| *One-legged endurance variables* | | | | |
| Peak work rate one-legged cycling | 62 ± 19 | 122 ± 30 | -67 (-58, -77) | < 0.001 # |
| Peak work rate one-legged cycling/peak work rate two-legged cycling (%) | 68 ± 13 | 63 ± 7 | 6 (11, 1) | 0.020 # |
| *Muscle strength and torque* | | | | |
| 1RM knee extension (kg) | 17 ± 7 | 22 ± 9 | -7 (-5, -9) | < 0.001 # |
| 1RM leg press (kg) | 104 ± 35 | 135 ± 29 | -36 (-26, -47) | < 0.001 # |
| 1RM chest press (kg) | 45 ± 16 | 53 ± 18 | -13 (-8, -18) | < 0.001 # |
| Peak torque (Nm), 60° · sec$^{-1}$ | 110 ± 38 | 122 ± 36 | -27 (-17, -36) | < 0.001 # |
| Peak torque (Nm), 180° · sec$^{-1}$ | 70 ± 28 | 76 ± 25 | -17 (-9, -24) | < 0.001 # |
| Peak torque (Nm), 240° · sec$^{-1}$ | 57 ± 23 | 62 ± 21 | -15 (-9, -20) | < 0.001 # |
| Handgrip strength (kg) | 42 ± 13 | 41 ± 11 | -3 (1, -6) | 0.112 |
| *Muscle thickness (mm)* | | | | |
| M. vastus lateralis | 19 ± 4 | 21 ± 3 | -2 (-1, -3) | 0.002 # |
| M. rectus femoris | 12 ± 4 | 15 ± 4 | -4 (-2, -5) | < 0.001 # |
| *M. vastus lateralis characteristics* | | | | |
| Cross-sectional area, µm$^2$ | | | | |
| Type I | 4614 ± 1088 | 3720 ± 951 | 449 (827, 70) | 0.020 # |
| Type II | 3639 ± 1235 | 3059 ± 1121 | 182 (482, -118) | 0.232 |
| Type I+II | 3978 ± 1066 | 3352 ± 943 | 184 (478, -109) | 0.216 |
| Myonuclei · fiber$^{-1}$ | | | | |
| Type I | 2.2 ± 0.9 | 2.1 ± 0.9 | -0.1 (0.2, -0.4) | 0.357 |
| Type II | 2.1 ± 0.7 | 1.9 ± 0.7 | -0.1 (0.2, -0.3) | 0.504 |
| Type I+II | 2.2 ± 0.7 | 2.0 ± 0.6 | -0.1 (0.2, -0.3) | 0.513 |
| Myonuclei domain, mean cross sectional area/nuclei *per* fiber, µm$^2$ | | | | |
| Type I | 2292 ± 585 | 1928 ± 1030 | 360 (613, 107) | 0.006 # |
| Type II | 1775 ± 529 | 1740 ± 1049 | -62 (191, -316) | 0.628 |
| Type I+II | 1835 ± 371 | 1771 ± 739 | 9 (244, -226) | 0.940 |

| | | | | | |
|---|---|---|---|---|---|
| | Fiber type proportion, % | | | | |
| | Type I | 52 ± 15 | 65 ± 14 | -16 (-9, -24) | < 0.001 # |
| | Type IIA | 32 ± 12 | 23 ± 11 | 10 (16, 4) | 0.001 # |
| | Type IIX | 13 ± 7 | 9 ± 6 | 5 (9, 1) | 0.007 # |
| | Type IIA/IIX | 3 ± 2 | 2 ± 2 | 0.7 (1.9, -0.4) | 0.159 |
| | Total RNA, ng · $ml^{-1}$ | 403 ± 86 | 432 ± 92 | -24 (10, -57) | 0.168 |
| *Habitual dietary data* $ | | | | | |
| | Kilocalories · $day^{-1}$ | 1768 ± 576 | 1937 ± 585 | -268 (34, -570) | 0.081 |
| | Protein (gram · $kg^{-1}$ · $day^{-1}$) | 1.2 ± 0.3 | 1.3 ± 0.4 | -0.1 (0.1, -0.3) | 0.458 |
| | Fat (gram · $kg^{-1}$ · $day^{-1}$) | 1.0 ± 0.4 | 1.0 ± 0.4 | -0.1 (0.2, -0.3) | 0.544 |
| | Carbohydrates (gram · $kg^{-1}$ · $day^{-1}$) | 2.6 ± 0.9 | 2.7 ± 1.1 | -0.2 (0.4, -0.8) | 0.539 |
| | Alcohol (units · $day^{-1}$) | 0.5 ± 0.8 | 0.8 ± 1.0 | -0.4 (0.1, -1.0) | 0.118 |

[£], as defined by Baumgartner *et al*.[E12]; [$], dietary registration was performed in the latter part of the training intervention (see Figure 2); [#], significant difference between COPD and Healthy (alpha level, $p$=0.05). Baseline values for each study cluster are presented as means ± standard deviations. Estimated differences between study groups are presented as means with 95% confidence limits.

**Table E3.** Genes identified as differentially expressed at baseline between COPD and Healthy in genome-wide transcriptome analyses (RNA-seq). RNA-seq analyses were performed as previously described.[E1,4]

| Ensembl gene ID | Gene Symbol | Log fold-change | SE | *Z*-value | *P*-value | Adjusted *P*-value[a] |
|---|---|---|---|---|---|---|
| ENSG00000146416 | AIG1 | -0.48 | 0.08 | -6.025 | 1.69e-09 | 2.56e-05 |
| ENSG00000112796 | ENPP5 | -0.57 | 0.10 | -5.556 | 2.75e-08 | 5.81e-05 |
| ENSG00000137942 | FNBP1L | -0.37 | 0.07 | -5.537 | 3.08e-08 | 5.81e-05 |
| ENSG00000143507 | DUSP10 | 0.44 | 0.08 | 5.612 | 2.00e-08 | 5.81e-05 |
| ENSG00000146477 | SLC22A3 | 0.88 | 0.16 | 5.555 | 2.78e-08 | 5.81e-05 |
| ENSG00000152782 | PANK1 | -0.44 | 0.08 | -5.601 | 2.14e-08 | 5.81e-05 |
| ENSG00000189067 | LITAF | 0.56 | 0.10 | 5.585 | 2.34e-08 | 5.81e-05 |
| ENSG00000205678 | TECRL | -0.67 | 0.12 | -5.620 | 1.91e-08 | 5.81e-05 |
| ENSG00000102007 | PLP2 | 0.50 | 0.09 | 5.495 | 3.91e-08 | 5.90e-05 |
| ENSG00000133816 | MICAL2 | 0.44 | 0.08 | 5.478 | 4.31e-08 | 5.91e-05 |
| | MICALCL | 0.44 | 0.08 | 5.478 | 4.31e-08 | 5.91e-05 |
| ENSG00000120658 | ENOX1 | 0.80 | 0.15 | 5.397 | 6.78e-08 | 8.16e-05 |
| ENSG00000150722 | PPP1R1C | -0.71 | 0.13 | -5.391 | 7.02e-08 | 8.16e-05 |
| ENSG00000113448 | PDE4D | 0.42 | 0.08 | 5.355 | 8.55e-08 | 9.22e-05 |
| ENSG00000048052 | HDAC9 | -0.59 | 0.11 | -5.242 | 1.59e-07 | 1.26e-04 |
| ENSG00000105835 | NAMPT | -0.38 | 0.07 | -5.253 | 1.50e-07 | 1.26e-04 |
| ENSG00000136040 | PLXNC1 | -0.52 | 0.10 | -5.251 | 1.51e-07 | 1.26e-04 |
| ENSG00000073910 | FRY | -0.43 | 0.08 | -5.225 | 1.74e-07 | 1.31e-04 |
| ENSG00000151746 | BICD1 | -0.61 | 0.12 | -5.172 | 2.31e-07 | 1.65e-04 |
| ENSG00000267296 | CEBPA-DT | 0.56 | 0.11 | 5.165 | 2.40e-07 | 1.65e-04 |
| ENSG00000225549 | Not mapped[d] | -0.92 | 0.18 | -5.146 | 2.66e-07 | 1.73e-04 |
| ENSG00000198729 | PPP1R14C | 0.44 | 0.09 | 5.126 | 2.96e-07 | 1.79e-04 |
| ENSG00000237301 | Not mapped[d] | 0.92 | 0.18 | 5.095 | 3.48e-07 | 1.95e-04 |
| ENSG00000091879 | ANGPT2 | 0.65 | 0.13 | 4.990 | 6.04e-07 | 2.95e-04 |
| ENSG00000151276 | MAGI1 | -0.36 | 0.07 | -4.994 | 5.90e-07 | 2.95e-04 |
| ENSG00000196152 | ZNF79 | 0.43 | 0.09 | 4.989 | 6.06e-07 | 2.95e-04 |
| ENSG00000183625 | CCR3 | -0.96 | 0.20 | -4.927 | 8.37e-07 | 3.83e-04 |
| ENSG00000140416 | TPM1 | 0.46 | 0.09 | 4.871 | 1.11e-06 | 4.78e-04 |
| ENSG00000130595 | TNNT3 | 0.41 | 0.08 | 4.856 | 1.20e-06 | 5.02e-04 |
| ENSG00000186352 | ANKRD37 | 0.59 | 0.12 | 4.849 | 1.24e-06 | 5.07e-04 |
| ENSG00000099194 | SCD | 1.04 | 0.22 | 4.797 | 1.61e-06 | 6.40e-04 |
| ENSG00000107282 | APBA1 | -0.43 | 0.09 | -4.768 | 1.86e-06 | 7.20e-04 |
| ENSG00000154814 | OXNAD1 | -0.40 | 0.08 | -4.762 | 1.92e-06 | 7.25e-04 |
| ENSG00000132953 | XPO4 | -0.54 | 0.11 | -4.727 | 2.28e-06 | 7.82e-04 |
| ENSG00000123700 | KCNJ2 | 0.42 | 0.09 | 4.668 | 3.04e-06 | 9.50e-04 |
| ENSG00000133794 | ARNTL | 0.54 | 0.12 | 4.665 | 3.09e-06 | 9.50e-04 |
| ENSG00000164197 | RNF180 | -0.35 | 0.08 | -4.616 | 3.91e-06 | 0.001 |
| ENSG00000144668 | ITGA9 | 0.38 | 0.08 | 4.611 | 4.01e-06 | 0.001 |
| ENSG00000137804 | NUSAP1 | 0.37 | 0.08 | 4.601 | 4.20e-06 | 0.001 |
| ENSG00000143549 | TPM3 | -0.44 | 0.10 | -4.552 | 5.32e-06 | 0.001 |
| ENSG00000226306 | NPY6R | -0.52 | 0.11 | -4.548 | 5.41e-06 | 0.001 |
| ENSG00000116741 | RGS2 | 0.70 | 0.15 | 4.544 | 5.51e-06 | 0.001 |
| ENSG00000159884 | CCDC107 | 0.43 | 0.10 | 4.538 | 5.68e-06 | 0.001 |
| ENSG00000184588 | PDE4B | 0.45 | 0.10 | 4.521 | 6.16e-06 | 0.002 |
| ENSG00000134986 | NREP | -0.48 | 0.11 | -4.513 | 6.39e-06 | 0.002 |
| ENSG00000105612 | DNASE2 | 0.51 | 0.11 | 4.499 | 6.84e-06 | 0.002 |
| ENSG00000066382 | MPPED2 | -0.48 | 0.11 | -4.489 | 7.15e-06 | 0.002 |
| ENSG00000147010 | SH3KBP1 | -0.36 | 0.08 | -4.469 | 7.85e-06 | 0.002 |
| ENSG00000108342 | CSF3 | -1.16 | 0.26 | -4.407 | 1.05e-05 | 0.002 |
| ENSG00000138061 | CYP1B1 | 0.47 | 0.11 | 4.404 | 1.06e-05 | 0.002 |
| ENSG00000162493 | PDPN | 0.35 | 0.08 | 4.408 | 1.04e-05 | 0.002 |

| Ensembl gene ID | Gene Symbol | Log fold-change | SE | *Z*-value | *P*-value | Adjusted *P*-value[a] |
|---|---|---|---|---|---|---|
| ENSG00000196526 | AFAP1 | 0.51 | 0.12 | 4.416 | 1.01e-05 | 0.002 |
| ENSG00000225613 | Not mapped[d] | 1.14 | 0.26 | 4.407 | 1.05e-05 | 0.002 |
| ENSG00000249464 | LINC01091 | 0.67 | 0.15 | 4.398 | 1.09e-05 | 0.002 |
| ENSG00000139998 | RAB15 | 0.53 | 0.12 | 4.385 | 1.16e-05 | 0.002 |
| ENSG00000138688 | KIAA1109 | -0.36 | 0.08 | -4.362 | 1.29e-05 | 0.002 |
| ENSG00000174437 | ATP2A2 | -0.44 | 0.10 | -4.361 | 1.30e-05 | 0.002 |
| ENSG00000119771 | KLHL29 | 0.53 | 0.12 | 4.351 | 1.35e-05 | 0.002 |
| ENSG00000134569 | LRP4 | 0.41 | 0.09 | 4.350 | 1.36e-05 | 0.002 |
| ENSG00000182985 | CADM1 | -0.35 | 0.08 | -4.352 | 1.35e-05 | 0.002 |
| ENSG00000139209 | SLC38A4 | 0.49 | 0.11 | 4.344 | 1.40e-05 | 0.002 |
| ENSG00000079156 | OSBPL6 | 0.37 | 0.08 | 4.340 | 1.42e-05 | 0.002 |
| ENSG00000077150 | NFKB2 | 0.42 | 0.10 | 4.333 | 1.47e-05 | 0.002 |
| ENSG00000163071 | SPATA18 | 0.52 | 0.12 | 4.323 | 1.54e-05 | 0.003 |
| ENSG00000180209 | MYLPF | 0.44 | 0.10 | 4.315 | 1.59e-05 | 0.003 |
| ENSG00000108960 | MMD | 0.35 | 0.08 | 4.302 | 1.69e-05 | 0.003 |
| ENSG00000176909 | MAMSTR | 0.52 | 0.12 | 4.297 | 1.73e-05 | 0.003 |
| ENSG00000138759 | FRAS1 | -0.37 | 0.09 | -4.251 | 2.13e-05 | 0.003 |
| ENSG00000186047 | DLEU7 | 0.93 | 0.22 | 4.249 | 2.15e-05 | 0.003 |
| | DLEU1-AS1 | 0.93 | 0.22 | 4.249 | 2.15e-05 | 0.003 |
| ENSG00000164649 | CDCA7L | -0.44 | 0.10 | -4.239 | 2.25e-05 | 0.003 |
| ENSG00000156265 | MAP3K7CL | 0.48 | 0.11 | 4.221 | 2.43e-05 | 0.003 |
| ENSG00000060656 | PTPRU | 0.52 | 0.12 | 4.214 | 2.51e-05 | 0.003 |
| ENSG00000162552 | WNT4 | 0.72 | 0.17 | 4.193 | 2.76e-05 | 0.004 |
| ENSG00000197442 | MAP3K5 | -0.40 | 0.09 | -4.190 | 2.78e-05 | 0.004 |
| ENSG00000223749 | Not mapped[d] | 1.35 | 0.32 | 4.187 | 2.82e-05 | 0.004 |
| ENSG00000175567 | UCP2 | 0.44 | 0.11 | 4.154 | 3.27e-05 | 0.004 |
| ENSG00000087903 | RFX2 | 0.56 | 0.13 | 4.135 | 3.55e-05 | 0.004 |
| ENSG00000138411 | HECW2 | -0.50 | 0.12 | -4.134 | 3.57e-05 | 0.004 |
| ENSG00000233621 | LINC01137 | 0.67 | 0.16 | 4.137 | 3.52e-05 | 0.004 |
| ENSG00000260337 | Not mapped[d] | 0.76 | 0.18 | 4.133 | 3.57e-05 | 0.004 |
| ENSG00000163823 | CCR1 | -0.65 | 0.16 | -4.127 | 3.67e-05 | 0.004 |
| ENSG00000106070 | GRB10 | -0.39 | 0.09 | -4.121 | 3.77e-05 | 0.004 |
| ENSG00000174791 | RIN1 | 0.96 | 0.23 | 4.108 | 3.99e-05 | 0.005 |
| ENSG00000196440 | ARMCX4 | 0.40 | 0.10 | 4.105 | 4.05e-05 | 0.005 |
| ENSG00000111602 | TIMELESS | 0.39 | 0.10 | 4.100 | 4.14e-05 | 0.005 |
| ENSG00000144908 | ALDH1L1 | 0.42 | 0.10 | 4.095 | 4.22e-05 | 0.005 |
| ENSG00000166833 | NAV2 | -0.40 | 0.10 | -4.093 | 4.25e-05 | 0.005 |
| ENSG00000101306 | MYLK2 | 0.35 | 0.09 | 4.079 | 4.51e-05 | 0.005 |
| ENSG00000285820 | Not mapped[d] | 1.43 | 0.35 | 4.076 | 4.58e-05 | 0.005 |
| ENSG00000129910 | CDH15 | 0.35 | 0.09 | 3.984 | 6.76e-05 | 0.007 |
| ENSG00000254901 | BORCS8 | 0.37 | 0.09 | 3.975 | 7.05e-05 | 0.007 |
| ENSG00000158486 | DNAH3 | -0.84 | 0.22 | -3.922 | 8.79e-05 | 0.008 |
| ENSG00000260391 | Not mapped[d] | 1.47 | 0.37 | 3.921 | 8.82e-05 | 0.008 |
| ENSG00000105327 | BBC3 | 0.72 | 0.19 | 3.903 | 9.50e-05 | 0.009 |
| ENSG00000183010 | PYCR1 | 0.66 | 0.17 | 3.898 | 9.69e-05 | 0.009 |
| ENSG00000226833 | LOC100505774 | -0.51 | 0.13 | -3.892 | 9.93e-05 | 0.009 |
| | LOC112267877 | -0.51 | 0.13 | -3.892 | 9.93e-05 | 0.009 |
| ENSG00000109061 | MYH1 | 0.68 | 0.18 | 3.889 | 1.01e-04 | 0.009 |
| ENSG00000089101 | CFAP61 | 0.52 | 0.13 | 3.878 | 1.05e-04 | 0.009 |
| ENSG00000168334 | XIRP1 | 0.42 | 0.11 | 3.857 | 1.15e-04 | 0.010 |
| ENSG00000178752 | ERFE | 0.83 | 0.21 | 3.851 | 1.17e-04 | 0.010 |
| ENSG00000272734 | Not mapped[d] | 0.43 | 0.11 | 3.853 | 1.17e-04 | 0.010 |
| ENSG00000105339 | DENND3 | -0.35 | 0.09 | -3.847 | 1.20e-04 | 0.010 |
| ENSG00000115129 | TP53I3 | 0.65 | 0.17 | 3.837 | 1.24e-04 | 0.010 |
| ENSG00000169710 | FASN | 0.78 | 0.20 | 3.838 | 1.24e-04 | 0.010 |
| ENSG00000169515 | CCDC8 | 0.72 | 0.19 | 3.827 | 1.30e-04 | 0.010 |

| Ensembl gene ID | Gene Symbol | Log fold-change | SE | *Z*-value | *P*-value | Adjusted *P*-value[a] |
|---|---|---|---|---|---|---|
| ENSG00000176749 | CDK5R1 | 0.40 | 0.11 | 3.818 | 1.35e-04 | 0.010 |
| ENSG00000109771 | LRP2BP | 0.44 | 0.12 | 3.812 | 1.38e-04 | 0.011 |
| ENSG00000068724 | TTC7A | 0.43 | 0.11 | 3.809 | 1.40e-04 | 0.011 |
| ENSG00000138615 | CILP | 0.40 | 0.11 | 3.806 | 1.41e-04 | 0.011 |
| ENSG00000109321 | AREG | 1.12 | 0.30 | 3.799 | 1.46e-04 | 0.011 |
| ENSG00000157330 | C1orf158 | 1.58 | 0.42 | 3.793 | 1.49e-04 | 0.011 |
| ENSG00000196296 | ATP2A1 | 0.43 | 0.11 | 3.796 | 1.47e-04 | 0.011 |
| ENSG00000228526 | MIR34AHG | 0.48 | 0.13 | 3.792 | 1.49e-04 | 0.011 |
| ENSG00000161513 | FDXR | 0.62 | 0.16 | 3.784 | 1.54e-04 | 0.011 |
| ENSG00000174032 | SLC25A30 | -0.39 | 0.10 | -3.775 | 1.60e-04 | 0.011 |
| ENSG00000104147 | OIP5 | 0.50 | 0.13 | 3.773 | 1.61e-04 | 0.011 |
| ENSG00000205106 | LINC02716 | 0.59 | 0.16 | 3.772 | 1.62e-04 | 0.011 |
| ENSG00000099999 | RNF215 | 0.42 | 0.11 | 3.760 | 1.70e-04 | 0.012 |
| ENSG00000196482 | ESRRG | -0.38 | 0.10 | -3.731 | 1.91e-04 | 0.013 |
| ENSG00000267080 | ASB16-AS1 | 0.36 | 0.10 | 3.713 | 2.05e-04 | 0.014 |
| ENSG00000205959 | Not mapped[d] | 0.39 | 0.11 | 3.684 | 2.30e-04 | 0.015 |
| ENSG00000138835 | RGS3 | -0.53 | 0.14 | -3.674 | 2.39e-04 | 0.015 |
| ENSG00000184545 | DUSP8 | 0.46 | 0.12 | 3.674 | 2.39e-04 | 0.015 |
| ENSG00000137193 | PIM1 | 0.46 | 0.13 | 3.669 | 2.43e-04 | 0.015 |
| ENSG00000262468 | Not mapped[d] | 0.51 | 0.14 | 3.665 | 2.47e-04 | 0.015 |
| ENSG00000023171 | GRAMD1B | 0.44 | 0.12 | 3.661 | 2.51e-04 | 0.015 |
| ENSG00000146166 | LGSN | -1.09 | 0.30 | -3.658 | 2.54e-04 | 0.015 |
| ENSG00000147256 | ARHGAP36 | 0.78 | 0.21 | 3.652 | 2.60e-04 | 0.016 |
| ENSG00000159259 | CHAF1B | 0.36 | 0.10 | 3.653 | 2.59e-04 | 0.016 |
| ENSG00000124587 | PEX6 | 0.44 | 0.12 | 3.634 | 2.79e-04 | 0.016 |
| ENSG00000215018 | COL28A1 | 0.35 | 0.10 | 3.607 | 3.10e-04 | 0.017 |
| ENSG00000139292 | LGR5 | -0.49 | 0.14 | -3.595 | 3.25e-04 | 0.018 |
| ENSG00000099308 | MAST3 | 0.66 | 0.18 | 3.589 | 3.32e-04 | 0.018 |
| ENSG00000102468 | HTR2A | -0.81 | 0.23 | -3.589 | 3.32e-04 | 0.018 |
| ENSG00000110660 | SLC35F2 | 0.54 | 0.15 | 3.586 | 3.36e-04 | 0.018 |
| ENSG00000089847 | ANKRD24 | 0.70 | 0.19 | 3.583 | 3.40e-04 | 0.018 |
| ENSG00000118515 | SGK1 | 0.44 | 0.12 | 3.583 | 3.40e-04 | 0.018 |
| ENSG00000124935 | SCGB1D2 | -0.74 | 0.21 | -3.556 | 3.76e-04 | 0.020 |
| ENSG00000163492 | CCDC141 | -0.44 | 0.12 | -3.553 | 3.81e-04 | 0.020 |
| ENSG00000184349 | EFNA5 | 0.60 | 0.17 | 3.551 | 3.84e-04 | 0.020 |
| ENSG00000064655 | EYA2 | 0.60 | 0.17 | 3.541 | 3.99e-04 | 0.020 |
| ENSG00000091513 | TF | 0.43 | 0.12 | 3.540 | 4.00e-04 | 0.020 |
| ENSG00000138379 | MSTN | 0.47 | 0.13 | 3.544 | 3.94e-04 | 0.020 |
| ENSG00000184347 | SLIT3 | 0.36 | 0.10 | 3.533 | 4.11e-04 | 0.020 |
| ENSG00000235070 | Not mapped[d] | -0.59 | 0.17 | -3.528 | 4.19e-04 | 0.021 |
| ENSG00000163879 | DNALI1 | 0.39 | 0.11 | 3.518 | 4.36e-04 | 0.021 |
| ENSG00000119969 | HELLS | 0.53 | 0.15 | 3.505 | 4.57e-04 | 0.022 |
| ENSG00000175489 | LRRC25 | -0.57 | 0.16 | -3.495 | 4.74e-04 | 0.022 |
| ENSG00000185105 | MYADML2 | 0.36 | 0.10 | 3.492 | 4.79e-04 | 0.023 |
| ENSG00000104313 | EYA1 | -0.42 | 0.12 | -3.489 | 4.85e-04 | 0.023 |
| ENSG00000258647 | Not mapped[d] | 0.74 | 0.21 | 3.483 | 4.96e-04 | 0.023 |
| ENSG00000260604 | Not mapped[d] | -0.63 | 0.18 | -3.484 | 4.94e-04 | 0.023 |
| ENSG00000278464 | Not mapped[d] | 0.43 | 0.12 | 3.483 | 4.95e-04 | 0.023 |
| ENSG00000075240 | GRAMD4 | 0.37 | 0.11 | 3.472 | 5.16e-04 | 0.023 |
| ENSG00000086967 | MYBPC2 | 0.41 | 0.12 | 3.473 | 5.15e-04 | 0.023 |
| ENSG00000145626 | UGT3A1 | 0.41 | 0.12 | 3.477 | 5.07e-04 | 0.023 |
| ENSG00000161036 | LRWD1 | 0.51 | 0.15 | 3.471 | 5.19e-04 | 0.023 |
| ENSG00000212907 | ND4L | -0.35 | 0.10 | -3.464 | 5.31e-04 | 0.023 |
| ENSG00000198915 | RASGEF1A | -0.62 | 0.18 | -3.459 | 5.41e-04 | 0.023 |
| ENSG00000106992 | AK1 | 0.40 | 0.12 | 3.454 | 5.53e-04 | 0.024 |
| ENSG00000277758 | LOC102724488 | 0.89 | 0.26 | 3.432 | 6.00e-04 | 0.025 |

| Ensembl gene ID | Gene Symbol | Log fold-change | SE | *Z*-value | *P*-value | Adjusted *P*-value[a] |
|---|---|---|---|---|---|---|
| ENSG00000197361 | FBXL22 | 0.49 | 0.14 | 3.404 | 6.63e-04 | 0.027 |
| ENSG00000231607 | DLEU2 | 0.35 | 0.10 | 3.393 | 6.90e-04 | 0.028 |
| ENSG00000158008 | EXTL1 | -0.58 | 0.17 | -3.392 | 6.94e-04 | 0.028 |
| ENSG00000140798 | ABCC12 | -0.70 | 0.21 | -3.389 | 7.01e-04 | 0.028 |
| ENSG00000165887 | ANKRD2 | 0.69 | 0.21 | 3.383 | 7.16e-04 | 0.028 |
| ENSG00000105877 | DNAH11 | 0.98 | 0.29 | 3.383 | 7.18e-04 | 0.028 |
| ENSG00000156463 | SH3RF2 | 0.40 | 0.12 | 3.373 | 7.45e-04 | 0.028 |
| ENSG00000285155 | Not mapped[d] | -0.39 | 0.11 | -3.372 | 7.45e-04 | 0.028 |
| ENSG00000168528 | SERINC2 | 0.51 | 0.15 | 3.366 | 7.62e-04 | 0.029 |
| ENSG00000188488 | SERPINA5 | -0.69 | 0.21 | -3.356 | 7.90e-04 | 0.030 |
| ENSG00000125844 | RRBP1 | 0.36 | 0.11 | 3.348 | 8.13e-04 | 0.030 |
| ENSG00000108932 | SLC16A6 | 0.58 | 0.18 | 3.336 | 8.51e-04 | 0.031 |
| ENSG00000130600 | H19 | 0.56 | 0.17 | 3.330 | 8.67e-04 | 0.031 |
| ENSG00000154080 | CHST9 | -0.56 | 0.17 | -3.336 | 8.49e-04 | 0.031 |
| ENSG00000174996 | KLC2 | 0.39 | 0.12 | 3.331 | 8.66e-04 | 0.031 |
| ENSG00000188582 | PAQR9 | -0.47 | 0.14 | -3.337 | 8.48e-04 | 0.031 |
| ENSG00000284820 | Not mapped[d] | 0.61 | 0.18 | 3.331 | 8.65e-04 | 0.031 |
| ENSG00000171617 | ENC1 | 0.41 | 0.12 | 3.328 | 8.74e-04 | 0.031 |
| ENSG00000047662 | FAM184B | 0.72 | 0.22 | 3.316 | 9.12e-04 | 0.032 |
| ENSG00000172932 | ANKRD13D | 0.41 | 0.13 | 3.314 | 9.21e-04 | 0.032 |
| ENSG00000158458 | NRG2 | 0.63 | 0.19 | 3.313 | 9.24e-04 | 0.032 |
| ENSG00000279529 | Not mapped[d] | 0.43 | 0.13 | 3.309 | 9.37e-04 | 0.032 |
| ENSG00000284693 | LINC02606 | -0.48 | 0.14 | -3.311 | 9.29e-04 | 0.032 |
| ENSG00000140795 | MYLK3 | -0.35 | 0.11 | -3.303 | 9.57e-04 | 0.033 |
| ENSG00000146005 | PSD2 | 0.79 | 0.24 | 3.301 | 9.63e-04 | 0.033 |
| ENSG00000148671 | ADIRF | 0.60 | 0.18 | 3.301 | 9.63e-04 | 0.033 |
| ENSG00000111245 | MYL2 | -0.35 | 0.11 | -3.281 | 0.001 | 0.034 |
| ENSG00000176134 | Not mapped[d] | -0.42 | 0.13 | -3.274 | 0.001 | 0.035 |
| ENSG00000071564 | TCF3 | 0.38 | 0.12 | 3.272 | 0.001 | 0.035 |
| ENSG00000214942 | Not mapped[d] | -0.72 | 0.22 | -3.270 | 0.001 | 0.035 |
| ENSG00000005206 | SPPL2B | 0.37 | 0.11 | 3.262 | 0.001 | 0.036 |
| ENSG00000181418 | DDN | 0.74 | 0.23 | 3.253 | 0.001 | 0.037 |
| ENSG00000215187 | FAM166B | 0.44 | 0.14 | 3.254 | 0.001 | 0.037 |
| ENSG00000052749 | RRP12 | 0.42 | 0.13 | 3.252 | 0.001 | 0.037 |
| ENSG00000264343 | NOTCH2NLA | 0.47 | 0.14 | 3.244 | 0.001 | 0.037 |
| ENSG00000173546 | CSPG4 | 0.46 | 0.14 | 3.231 | 0.001 | 0.038 |
| ENSG00000177551 | NHLH2 | 1.15 | 0.36 | 3.230 | 0.001 | 0.038 |
| ENSG00000117707 | PROX1 | -0.36 | 0.11 | -3.227 | 0.001 | 0.039 |
| ENSG00000225472 | Not mapped[d] | -0.51 | 0.16 | -3.227 | 0.001 | 0.039 |
| ENSG00000159713 | TPPP3 | 0.55 | 0.17 | 3.226 | 0.001 | 0.039 |
| ENSG00000205279 | CTXN3 | -0.72 | 0.22 | -3.221 | 0.001 | 0.039 |
| ENSG00000255495 | Not mapped[d] | 0.40 | 0.12 | 3.220 | 0.001 | 0.039 |
| ENSG00000149090 | PAMR1 | 0.46 | 0.14 | 3.211 | 0.001 | 0.040 |
| ENSG00000124374 | PAIP2B | -0.35 | 0.11 | -3.193 | 0.001 | 0.041 |
| ENSG00000072310 | SREBF1 | 0.48 | 0.15 | 3.188 | 0.001 | 0.042 |
| ENSG00000104889 | RNASEH2A | 0.47 | 0.15 | 3.184 | 0.001 | 0.042 |
| ENSG00000238083 | LRRC37A2 | 0.36 | 0.11 | 3.185 | 0.001 | 0.042 |
| ENSG00000270021 | Not mapped[d] | 0.52 | 0.16 | 3.184 | 0.001 | 0.042 |
| ENSG00000185847 | LINC01405 | -0.41 | 0.13 | -3.177 | 0.001 | 0.043 |
| ENSG00000248587 | Not mapped[d] | 0.36 | 0.11 | 3.173 | 0.002 | 0.043 |
| ENSG00000105738 | SIPA1L3 | 0.37 | 0.12 | 3.161 | 0.002 | 0.044 |
| ENSG00000273301 | Not mapped[d] | -0.86 | 0.27 | -3.145 | 0.002 | 0.046 |
| ENSG00000077943 | ITGA8 | -0.41 | 0.13 | -3.136 | 0.002 | 0.047 |
| ENSG00000241288 | LINC02614 | 0.43 | 0.14 | 3.138 | 0.002 | 0.047 |
| ENSG00000127191 | TRAF2 | 0.52 | 0.17 | 3.134 | 0.002 | 0.047 |
| ENSG00000283563 | ZCWPW2 | -0.36 | 0.11 | -3.129 | 0.002 | 0.047 |

| **Ensembl gene ID** | **Gene Symbol** | **Log fold-change** | **SE** | ***Z*-value** | ***P*-value** | **Adjusted *P*-value[a]** |
|---|---|---|---|---|---|---|
| ENSG00000140280 | LYSMD2 | 0.37 | 0.12 | 3.127 | 0.002 | 0.047 |
| ENSG00000070601 | FRMPD1 | -0.36 | 0.12 | -3.117 | 0.002 | 0.048 |
| ENSG00000108231 | LGI1 | -0.37 | 0.12 | -3.112 | 0.002 | 0.049 |
| ENSG00000220563 | Not mapped[d] | 0.37 | 0.12 | 3.110 | 0.002 | 0.049 |
| ENSG00000250303 | LINC02762 | -0.37 | 0.12 | -3.109 | 0.002 | 0.049 |
| ENSG00000166123 | GPT2 | -0.37 | 0.12 | -3.107 | 0.002 | 0.049 |
| ENSG00000167037 | SGSM1 | -0.60 | 0.19 | -3.102 | 0.002 | 0.049 |
| ENSG00000153822 | KCNJ16 | 0.59 | 0.19 | 3.098 | 0.002 | 0.049 |

[a] *P*-values are adjusted for FDR. [b] Raw *P*-values from simulation based tests of uniformity of residuals where low values indicate problematic models. [d] No official gene symbol available, not included in enrichment analyses.

**Table E4.** Gene ontology (GO) analysis of genome-wide transcriptome data (RNA-seq; COPD vs. Healthy), performed as previously described.[E1,4]

| Comparison | Gene set category | Gene set | Significance category[a] | Set size[b] | Rank *P*-value[c] | % MSD > 0[d] | GSEA *P*-value[e] | NES | LE[f] | $Log_2$ Fold-change in LE [min, max] |
|---|---|---|---|---|---|---|---|---|---|---|
| Baseline: COPD vs. Healthy | Biological process | Actin filament based movement | Rank | 118 (153) | 4.47e-05 | 30.5% | 0.760 | 1.08 | 20 (85%) | 0.6 [0.29, 0.97] |
| | | Actin mediated cell contraction | Rank | 92 (123) | 4.29e-05 | 34.8% | 0.728 | 1.12 | 14 (92.9%) | 0.63 [0.41, 0.97] |
| | | Fatty acid metabolic process | Rank | 279 (396) | 8.48e-06 | 29.7% | 0.602 | -1.12 | 61 (83.6%) | -0.32 [-0.96, -0.15] |
| | | Monocarboxylic acid metabolic process | Rank | 469 (672) | 8.48e-06 | 29.4% | 0.468 | -1.17 | 72 (97.2%) | -0.36 [-1.17, -0.16] |
| | | Muscle contraction | Rank | 252 (362) | 4.27e-05 | 29.4% | 0.767 | 1.05 | 34 (82.4%) | 0.59 [0.25, 1.01] |
| | | Muscle filament sliding | Rank | 31 (39) | 1.39e-04 | 54.8% | 0.728 | 1.15 | 10 (90%) | 0.61 [0.29, 0.97] |
| | | Muscle system process | Rank | 321 (467) | 8.48e-06 | 29.9% | 0.740 | 1.08 | 45 (82.2%) | 0.56 [0.25, 1.03] |
| | Cellular component | Inner mitochondrial membrane protein complex | GSEA | 114 (138) | 0.771 | 27.2% | 0.003 | -1.83 | 39 (76.9%) | -0.22 [-0.37, -0.12] |
| | | Mitochondrial matrix | GSEA | 436 (473) | 0.122 | 29.4% | 2.19e-04 | -1.61 | 120 (88.3%) | -0.23 [-0.53, -0.12] |
| | | Mitochondrial protein complex | GSEA | 234 (265) | 0.933 | 26.1% | 3.66e-05 | -1.90 | 70 (80%) | -0.21 [-0.37, -0.1] |
| | | Organelle inner membrane | GSEA | 461 (549) | 0.826 | 25.4% | 0.005 | -1.43 | 92 (95.7%) | -0.24 [-0.56, -0.14] |
| | | Actin cytoskeleton | Rank | 392 (503) | 1.87e-04 | 28.1% | 0.304 | 1.29 | 93 (74.2%) | 0.41 [0.14, 0.98] |
| | | Contractile fiber | Rank | 191 (238) | 2.24e-05 | 33% | 0.505 | 1.21 | 49 (83.7%) | 0.41 [0.16, 1] |
| | Molecular function | G protein coupled receptor activity | GSEA | 146 (867) | 0.411 | 22.6% | 0.018 | -1.75 | 29 (69%) | -0.52 [-1.39, -0.17] |
| Post-RT (13 weeks training): ΔCOPD vs ΔHealthy | Biological process | Proteasomal protein catabolic process | Rank | 421 (481) | 0.019 | 29.2% | 0.591 | -1.08 | 97 (82.5%) | -0.43 [-1.26, -0.19] |
| | | Regulation of cholesterol efflux | Rank | 25 (46) | 0.019 | 48% | 0.102 | -1.50 | 13 (84.6%) | -0.54 [-1.32, -0.29] |
| | | Regulation of protein catabolic process | Rank | 327 (395) | 0.019 | 31.2% | 0.293 | -1.17 | 71 (97.2%) | -0.46 [-1.26, -0.23] |
| | Cellular component | Actin cytoskeleton | Consensus | 392 (503) | 0.002 | 29.1% | 5.68e-06 | -1.38 | 133 (75.2%) | -0.44 [-1.17, -0.16] |
| | | Actin filament bundle | Consensus | 67 (75) | 5.45e-04 | 38.8% | 0.016 | -1.47 | 31 (74.2%) | -0.48 [-1.17, -0.2] |
| | | Actomyosin | Consensus | 69 (78) | 4.62e-04 | 37.7% | 0.011 | -1.48 | 31 (74.2%) | -0.49 [-1.17, -0.2] |
| | | Contractile fiber | Consensus | 191 (238) | 1.04e-05 | 33.5% | 1.56e-04 | -1.44 | 63 (87.3%) | -0.47 [-1.17, -0.19] |
| | | I band | Consensus | 114 (140) | 4.71e-04 | 33.3% | 2.28e-04 | -1.52 | 40 (87.5%) | -0.48 [-1.17, -0.2] |
| | | Adherens junction | GSEA | 127 (166) | 0.244 | 27.6% | 0.005 | -1.42 | 44 (65.9%) | -0.47 [-1.17, -0.16] |
| | | Cell cell junction | GSEA | 344 (493) | 0.161 | 26.5% | 1.91e-04 | -1.34 | 116 (63.8%) | -0.43 [-1.17, -0.16] |
| | | Cell substrate junction | GSEA | 359 (423) | 0.305 | 27.9% | 0.003 | -1.31 | 112 (68.8%) | -0.43 [-0.96, -0.16] |
| | | Collagen containing extracellular matrix | GSEA | 214 (427) | 0.999 | 20.1% | 0.005 | -1.34 | 74 (51.4%) | -0.47 [-1.56, -0.19] |
| | | Extrinsic component of cytoplasmic side of plasma membrane | GSEA | 65 (99) | 0.305 | 24.6% | 0.003 | -1.56 | 15 (100%) | -0.53 [-0.89, -0.3] |
| | | Extrinsic component of plasma membrane | GSEA | 109 (172) | 0.458 | 22% | 0.005 | -1.45 | 25 (84%) | -0.51 [-0.89, -0.27] |
| | | Polymeric cytoskeletal fiber | GSEA | 437 (756) | 0.110 | 25.9% | 0.005 | -1.25 | 135 (71.1%) | -0.43 [-1.17, -0.17] |
| | | Heterochromatin | Rank | 63 (78) | 0.004 | 39.7% | 0.063 | -1.40 | 19 (94.7%) | -0.48 [-1.05, -0.17] |
| | Molecular function | Actin binding | Consensus | 336 (437) | 0.001 | 30.7% | 3.17e-07 | -1.42 | 125 (75.2%) | -0.44 [-1.17, -0.16] |
| | | Actin filament binding | Consensus | 162 (206) | 0.002 | 32.7% | 0.001 | -1.43 | 65 (70.8%) | -0.46 [-1.13, -0.21] |
| | | Chromatin binding | Consensus | 448 (596) | 0.001 | 29.2% | 0.025 | -1.23 | 94 (92.6%) | -0.44 [-1.05, -0.17] |
| | | Molecular adaptor activity | Consensus | 252 (314) | 0.001 | 30.6% | 0.048 | -1.25 | 80 (70%) | -0.44 [-1.14, -0.16] |
| | | Cell adhesion molecule binding | GSEA | 407 (544) | 0.384 | 25.3% | 5.69e-04 | -1.31 | 120 (75.8%) | -0.45 [-1.56, -0.16] |
| | | Protein kinase activity | GSEA | 449 (563) | 0.353 | 25.6% | 9.88e-04 | -1.29 | 102 (81.4%) | -0.49 [-1.29, -0.22] |
| | | Protein serine threonine kinase activity | GSEA | 361 (434) | 0.167 | 27.1% | 0.004 | -1.28 | 83 (84.3%) | -0.48 [-1.26, -0.22] |
| | | Glutamate receptor binding | Rank | 31 (46) | 0.016 | 54.8% | 0.071 | -1.49 | 16 (100%) | -0.41 [-0.69, -0.16] |
| | | Nuclear receptor binding | Rank | 83 (101) | 0.016 | 37.3% | 0.812 | -1.03 | 21 (90.5%) | -0.43 [-1.05, -0.16] |

| **Comparison** | **Gene set category** | **Gene set** | **Significance category[a]** | **Set size[b]** | **Rank *P*-value[c]** | **% MSD > 0[d]** | **GSEA *P*-value[e]** | **NES** | **LE[f]** | **$Log_2$ Fold-change in LE [min, max]** |
|---|---|---|---|---|---|---|---|---|---|---|
| | | Protein macromolecule adaptor activity | Rank | 200 (244) | 6.58e-04 | 33% | 0.070 | -1.28 | 71 (69%) | -0.44 [-1.14, -0.16] |
| | | Signaling adaptor activity | Rank | 54 (68) | 0.016 | 40.7% | 0.343 | -1.25 | 22 (77.3%) | -0.42 [-0.77, -0.18] |
| | | Signaling receptor complex adaptor activity | Rank | 32 (41) | 0.016 | 43.8% | 0.267 | -1.32 | 10 (100%) | -0.49 [-0.77, -0.27] |
| | | Structural constituent of muscle | Rank | 33 (43) | 0.016 | 42.4% | 0.073 | -1.44 | 13 (92.3%) | -0.49 [-0.97, -0.26] |
| | | Ubiquitin binding | Rank | 71 (76) | 0.018 | 38% | 0.145 | -1.37 | 24 (91.7%) | -0.42 [-0.88, -0.25] |

[a] Consensus significance indicates agreement between directional (GSEA) and non-directional (Rank) hypothesis test of overrepresentation (see methods for details). [b] Indicates number of identified genes in the gene set and total number of genes in the gene set in parentheses. [c] Rank-based enrichment test, based on minimum significant difference (MSD), identifies gene sets that are overrepresented among top-ranked genes without a directional hypothesis. [d] Fraction of genes in gene set with unadjusted 95% CI not spanning zero, i.e. MSD > 0. [e] Gene-set enrichment analysis (GSEA) tests for overrepresentation among top and bottom genes based on $Log_2$ fold differences or changes × $-log_{10}$(*P*-values) in comparing differences at baseline or changes from baseline between COPD and Healthy. Positive normalized enrichment score (NES) indicate gene sets with higher expression in COPD than Healthy; negative NES indicate gene sets with lower expression at respective time-points. [f] Number of genes in leading edge (LE, genes that contributes to the enrichment score) with the fraction of leading edge genes with unadjusted 95% CI not spanning zero.

E4. Mølmen KS, Hammarström D, Pedersen K, Lie ACL, Steile RB, Nygaard H, Khan Y, Hamarsland H, Koll L, Hanestadhaugen M, Eriksen AL, Grindaker E, Whist JE, Buck D, Ahmad R, Strand TA, Rønnestad BR, Ellefsen S. Vitamin D3 supplementation does not enhance the effects of resistance training in older adults. *J Cachexia Sarcopenia Muscle*. 2021;In Press.

E5. R Core Team. R: A Language and Environment for Statistical Computing. R Foundation for Statistical Computing, Vienna, Austria.

E6. Borg GA. Psychophysical bases of perceived exertion. *Med Sci Sports Exerc*. 1982;14(5):377-381.

E7. Nuzzo JL, Taylor JL, Gandevia SC. CORP: Measurement of upper and lower limb muscle strength and voluntary activation. *J Appl Physiol*. 2018;126(3):513-543.

E8. Hamarsland H, Johansen MK, Seeberg F, Brochmann M, Garthe I, Benestad HB, Raastad T. Native Whey Induces Similar Adaptation to Strength Training as Milk, despite Higher Levels of Leucine, in Elderly Individuals. *Nutrients*. 2019;11(9):2094.

E9. Häkkinen K, Kallinen M, Linnamo V, Pastinen U-M, Newton RU, Kraemer WJ. Neuromuscular adaptations during bilateral versus unilateral strength training in middle aged and elderly men and women. *Acta Physiol Scand*. 1996;158(1):77-88.

E10. Ahtiainen JP, Walker S, Peltonen H, Holviala J, Sillanpää E, Karavirta L, Sallinen J, Mikkola J, Valkeinen H, Mero A, Hulmi JJ, Häkkinen K. Heterogeneity in resistance training-induced muscle strength and mass responses in men and women of different ages. *Age (Omaha)*. 2016;38(1):1-13.

E11. Haun CT, Vann CG, Roberts BM, Vigotsky AD, Schoenfeld BJ, Roberts MD. A critical evaluation of the biological construct skeletal muscle hypertrophy: Size matters but so does the measurement. *Front Physiol*. 2019;10(MAR):1-23.

E12. Baumgartner RN, Koehler KM, Gallagher D, Romero L, Heymsfield SB, Ross RR, Garry PJ, Lindeman RD. Epidemiology of sarcopenia among the elderly in New Mexico. *Am J Epidemiol*. 1998;147(8):755-763.
